# Culture-free *Helicobacter pylori* genomics reveals within-host virulence heterogeneity and antibiotic resistance

**DOI:** 10.64898/2026.10.23.26365955

**Authors:** Yujie Liu, Mi Ni, Xue-Song Zhang, Jessica Bass, Panchajanya Deshpande, Judith Romero-Gallo, Hao Chen, Yu Fan, Difei Wang, Wanjin Qiao, Yanchun Zhang, Edward A. Mead, Emmanuel Dias-Neto, Alejandro Corvalan, Rachel Brody, M. Constanza Camargo, Wendy Szymczak, Amy D. Klion, Richard M. Peek, Martin J. Blaser, Gang Fang

## Abstract

*Helicobacter pylori* (Hp) infects over 40% of the world’s population and causes most cases of gastric cancer worldwide, yet fundamental questions about its within-host genetic diversity remain unresolved. Decades of work on Hp genomics depends on culturing clinical isolates, which can distort within-host genetic heterogeneity that informs Hp virulence and antimicrobial resistance. Culture-free alternatives have been limited by the extremely low Hp abundance in gastric tissue (usually 10^−3^ to 10^−7^ of total genomic DNA), making direct metagenomics prohibitively expensive and existing enrichment strategies insufficient for whole-genome recovery. Here we develop ultra-sensitive Hp sequencing (uHp-seq) for reconstructing Hp genomes directly from gastric biopsies. uHp-seq achieves a thousand-fold Hp DNA enrichment and recovers near-complete genomes across samples spanning orders of magnitude in Hp abundance. Notably, individual hosts may harbor multiple co-existing Hp strains or cagA EPIYA variants with different oncogenic potential. Resistance-associated variants for six commonly used antibiotics are resolved with greater accuracy than PCR-based and probe-based commercial panels. Applied to minimally invasive gastric string samples, uHp-seq delivers reliable resistance profiles without endoscopy, opening a path to genome-guided Hp eradication.

## Introduction

*Helicobacter pylori* (Hp) infects over 40% of the world’s population and is the leading infectious cause of gastric cancer^1^, the fifth most common cancer and the fifth leading cause of cancer death worldwide^2^. Hp strains differ markedly in their capacity to cause disease, driven by variation in key virulence loci: the *cag* pathogenicity island, *cagA* EPIYA configuration, *vacA* alleles, and adhesin repertoires that together determine the intensity of host interaction and the risk of malignant progression^3–5^. Identifying patients at highest risk therefore depends on knowing the virulence configuration of the strain they carry. Importantly, selecting an effective eradication regimen requires knowing the strain’s antibiotic resistance genotype^6–8^, as rising antimicrobial resistance has driven first-line treatment success below 90% in most regions, as low as 60% for certain antibiotics in certain regions^9–12^. Despite four decades of Hp genomic research, this information cannot be efficiently obtained for individual patients in clinical practice.

The barrier is technical rather than conceptual. Hp is exceptionally diverse and individual strains evolve within hosts over years^13^, making whole-genome resolution essential for reliable biological and clinical inference. Nearly all Hp genomic insight has come from culture of clinical isolates. However, culture introduces bias by favoring strains that grow better under laboratory conditions, collapses complex multi-strain infections to a single strain, misses single-nucleotide polymorphisms (SNPs) and structural variations (SVs), and misrepresents the genomic landscape of Hp as it exists at the gastric mucosa.

Culture-free alternatives have been attempted, but each is bounded by the fundamental challenge of the Hp scarcity of Hp in gastric tissue: Hp constitutes only 10^−3^ to 10^−7^ of total genomic DNA, with the vast majority derived from human cells^14,15^. Conventional shotgun metagenomics is cost prohibitive to achieve sufficient Hp coverage. Raman-spectra-based sorting and laser capture microdissection can recover Hp cells prior to sequencing but require specialized instrumentation, scale poorly to clinical throughput^16,17^. PCR-based clinical assays detect a handful of resistance hotspots and virulence markers but miss important variants because they cannot resolve full genetic variations that shapes infection outcome^18–21^. Although probe-based enrichment is a generally applicable approach, it is often limited by hybridization bias^22,23,24^.

Here we report uHp-seq, a culture-free workflow that reconstructs Hp genomes directly from host-dominated clinical specimens by coupling methylation-guided depletion of non-Hp DNA with ultra-low-input sequencing optimized for the trace quantities of the enriched Hp DNA that remain. uHp-seq achieves a thousand-fold enrichment and recovers near-complete genomes from gastric biopsies and minimally invasive gastric string samples across a wide range of Hp abundance. We show that culture systematically misrepresents within-host Hp heterogeneity, missing mixed infections, collapsing co-existing *cagA* EPIYA variants with distinct oncogenic potential, and flattening gastric regional Hp genomic structure. In contrast, uHp-seq resolves these heterogeneity. Genome-wide profiling of antibiotic resistance variants via uHp-seq outperforms PCR-based and probe-based commercial panels. Applied to gastric string samples from patients with refractory infections, uHp-seq delivers comprehensive antibiotic resistance profiles without endoscopy, establishing a practical route to genome-guided eradication therapy.

## Results

### uHp-seq enables robust culture-free recovery of near-complete Hp genomes

To highlight the challenge of recovering Hp genomes without culturing, we showed that Hp constitutes 10⁻³ to 10⁻ of total sequencing reads in gastric biopsies, with the remainder dominated by host DNA (>99.99%) and a complex non-Hp microbial background, consistent across both public metagenomes (**Extended Data Fig. 1a)**^14,15^ and our in-house gastric biopsy data (**Fig. 1a-b**). Direct shotgun metagenomics is prohibitive at this abundance: reaching ≥10× Hp coverage from a 10⁻ sample requires thousands of gigabases per sample.

**Figure 1.**
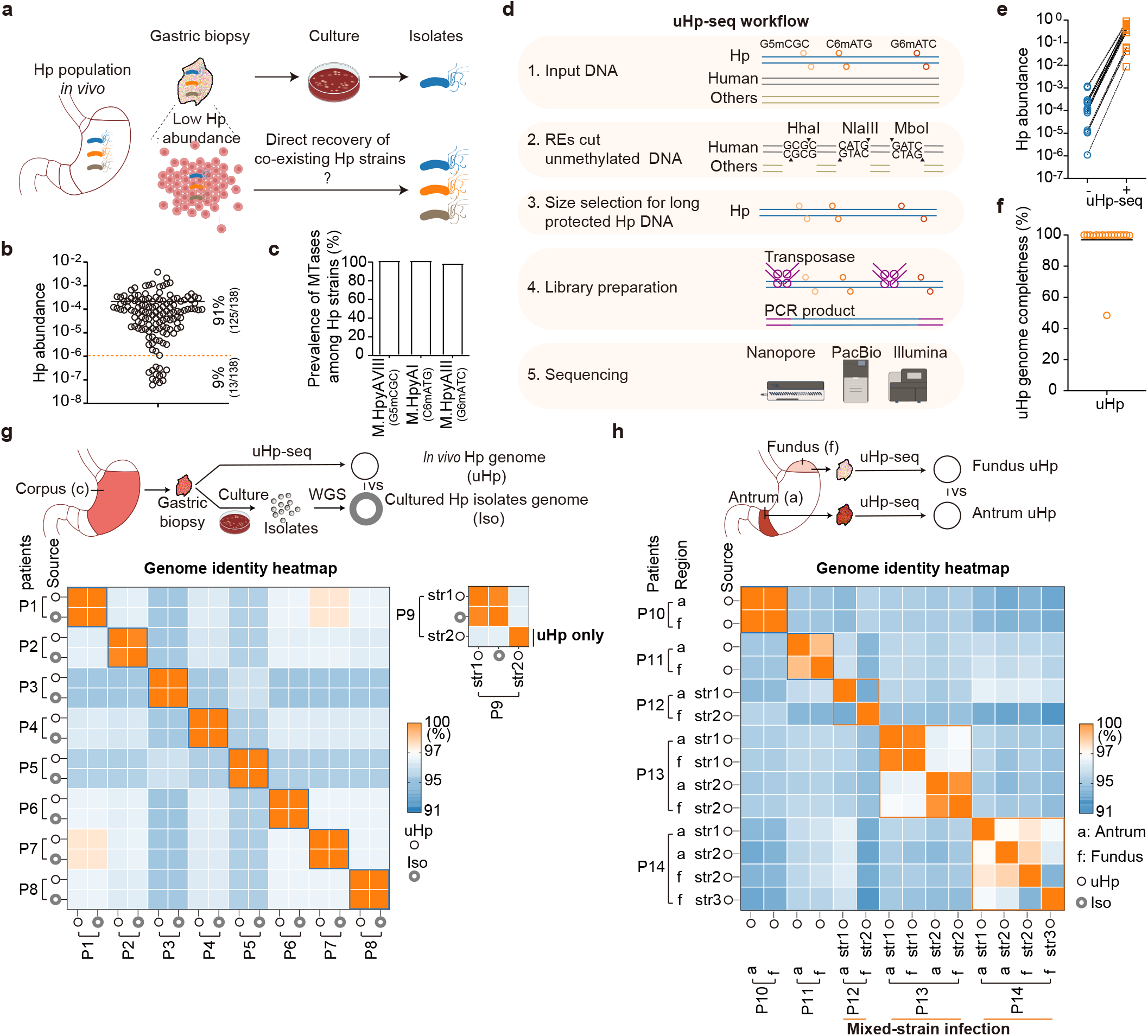
Development and validation of uHp-seq for culture-free recovery of complete Hp genomes from ultra-low-abundance gastric specimens. **a,** *In vivo* Hp population diversity may be distorted by culture, whereas direct shotgun metagenomic sequencing is limited by the extremely low abundance of Hp (10⁻³–10⁻) in gastric specimens dominated by human DNA (>99.99%). **b,** Hp abundance distribution in gastric biopsies, determined by qPCR (n=138). **c,** Prevalence of type II methyltransferases (MTases) across Hp genomes. Three conserved MTases are present in nearly all strains and form the basis of selective enrichment. **d,** Schematic of the uHp-seq workflow. Conserved Hp methylation motifs protect long bacterial DNA fragments from digestion by methylation-sensitive restriction endonucleases (REases), whereas unmethylated background DNA is fragmented. Protected fragments are enriched by size selection with high recovery (BluePippin), followed by transposase-based library preparation for ultra-low-input sequencing. **e**, uHp-seq effectively enriches Hp in gastric biopsy samples (n=17). **f,** uHp genome completeness estimated by mapping Hp reads to the genomes of their paired isolates (n=17). **g,** Validation of biopsy-derived genome reconstruction. Across eight matched biopsy–isolate pairs acquired from corpus biopsies, uHp-seq genomes (uHp) showed 99.9% nucleotide identity to paired cultured isolates (Iso). **h,** uHp-seq resolves intra-host Hp diversity across gastric regions. Antral and fundic biopsies frequently exhibited substantial genomic divergence, including mixed-strain infections and cases with up to three distinct strains detected within a single biopsy. The center line in each dot plot indicates the median.

We reasoned that a more effective solution would exploit a feature intrinsic to the Hp genome yet less dependent on specific sequences than PCR or probe-based approaches. Hp strains encode on average ∼20 DNA methyltransferases (MTases), three of which target highly conserved 4-mer motifs across nearly all clinical isolates: M.HpyAVIII (G5mCGC), M.HpyAI (C6mATG), and M.HpyAIII (G6mATC) (**Fig. 1c**, **Extended Data Fig. 1b**), as confirmed in more than 2,000 Hp strains using public datasets^25–27^ and *Helicobacter pylori* Genome Project (*Hp*GP) analysis^28^. These methylation marks shield Hp DNA from cleavage by motif-matched, methylation-sensitive restriction endonucleases (REases, HhaI, NlaIII, and MboI), while unmethylated host and non-Hp microbial DNA is digested^29^. Human DNA cutting and *in silico* digestion of ∼4,000 reference bacterial genomes confirmed that combined three-REase, HhaI, NlaIII, and MboI maximized cleavage of background DNA while preserving the largest fraction of Hp DNA as long fragments (**Extended Data Fig. 1c-d**).

Building on this principle, we developed uHp-seq (**Fig. 1d**). Genomic DNA from clinical specimens is co-digested with HhaI, NlaIII, and MboI, fragmenting unmethylated background DNA to <1.5 kb while leaving methylation-protected Hp DNA intact at >10 kb (**Extended Data Fig. 1d**). Long Hp fragments are then isolated by automated size selection on the BluePippin platform. Recovered Hp DNA is barcoded by transposase-mediated tagmentation using MuA transposase for nanopore sequencing (**Extended Data Fig. 1g-h**) or Tn5 for short-read sequencing (**Extended Data Fig. 2**).

To confirm that uHp-seq reconstructs the correct genome rather than a chimera or contaminant assembly, we applied uHp-seq to 17 gastric biopsy samples from nine patients and performed whole-genome sequencing (WGS) on paired cultured isolates from each using the same library preparation protocol, but without restriction enzyme digestion or size selection. uHp-seq increased the Hp abundance by over 1,000-20,000-folds (**Fig. 1e**), and the uHp reads achieved 100% genome completeness for 16 of the 17 samples when mapped to each of the corresponding paired isolate genomes (**Fig. 1f**). uHp genomes showed >99.9% average nucleotide identity (ANI) to their corresponding isolates (**Fig. 1g, Extended Data Fig.3a**). Importantly, the uHp-recovered genomes reveal greater genomic diversity than present in the paired isolates (**Extended Data Fig. 3b**). For example, patient P9 harbored a second distinct Hp strain detected in the biopsy but missed by culture (**Fig. 1g**).

Further applied at a larger scale to 94 gastric biopsies from five collections, including the 17 samples described above (**Supplementary Table 1**). uHp-seq delivered a median of ∼2,500-fold enrichment, significantly better than the microbial-enrichment method (MEM)^30^ and saponin-based enrichment^31^, two culture-free methods for enriching bacterial genomic DNA from tissues (**Extended Data Fig. 4a, b, Supplementary Table 2**). uHp-seq recovered Hp genomes including plasmids from 87 of the 94 samples (93%) with a high success rate to recover near-complete genomes (**Extended Data Fig. 4c, d**). Notably, uHp-seq recovered Hp genomes from all nine Hp-positive biopsy samples for which culture was not successful. uHp-seq consistently recovered genomes from samples with Hp abundance as low as 10⁻ (100 Hp genome copies in 200 ng of biopsy DNA), a threshold encompassing approximately 91% of gastric biopsy samples based on the observed abundance distribution (**Fig. 1b**). Reaching ≥10× genome coverage required only ∼0.2 Gb of sequencing per sample (**Extended Data Fig. 4e**), confirming compatibility with population-scale studies on standard sequencing budgets.

### uHp-seq reveals within-host Hp strain diversity and virulence heterogeneity

uHp-seq uncovered an additional co-existing Hp strain in patient P9 that had been missed by colony picking and culture (**Fig. 1g**), prompting a systematic characterization of within-host Hp heterogeneity across the broader samples. Among five patients with paired antrum and fundus biopsies, only one carried near-identical strains across both sites (patient P10, 99.9% ANI). The remaining four showed marked between-site divergence, including distinct strains in antrum versus fundus (patient P12) and mixed infections within a single biopsy (patients P13 and P14, with patient P14 carrying three distinct strains in one sample) (**Fig. 1h**). Antrum–fundus pairs showed greater divergence than antrum–corpus pairs (**Extended Data Fig. 5a-b**), consistent with stronger biogeographic structuring along the proximal–distal axis of the stomach. These findings indicate that Hp genomics of single-site, single-colony, or pooled colonies picked from the primary isolation plates underestimates within-host strain heterogeneity, and that uHp-seq, by recovering native (*in vivo*) genomes directly from tissue, provides the resolution needed to characterize it.

To define within-host Hp genomic heterogeneity at SNP resolution, we compared uHp-seq genomes against cultured isolates across samples with paired uHp-recovered genomes (single strain) and isolate genomes (**Fig. 2a, Supplementary Table 3**). SNPs present *in vivo* but absent from culture comprised ∼21,844 positions across the cohort, of which 7,550 were protein-changing (**Fig. 2b**). When mapped to reference strain Hp26695, most *in vivo* variants missed by culture were subject-specific, while 576 recurrent SNPs were detected across at least two patients (**Fig. 2c-d, Supplementary Tables 4** and **5**), including several genes associated with host interaction and gastric adaptation such as TonB-dependent transport genes (*frpB2* and *tonB2*), outer membrane proteins (*sabA*, *hopQ*, and *babA*), flagellar assembly (*fliF*), and established virulence determinants. uHp-seq also captured ∼686 SNPs that differ between gastric regions, with 10 recurrent variants detected across at least two patients (**Supplementary Table 6**, **Fig. 2b; Methods**).

**Figure 2.**
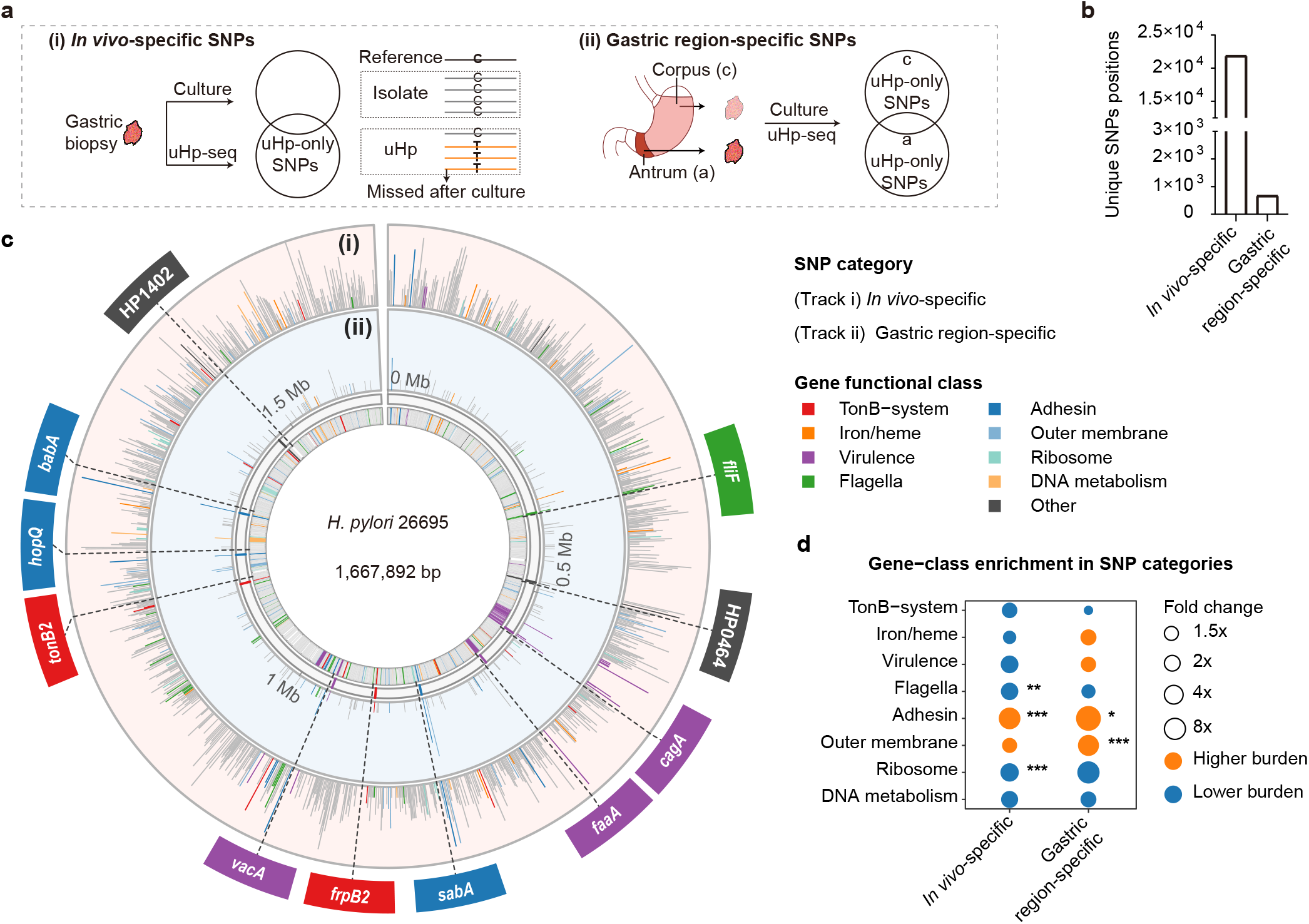
uHp-seq reveals Hp SNPs missed after culture. **a,** Study design for identifying uHp-specific SNPs across two perspectives. (i) *In vivo*-specific SNPs, identified by comparing paired uHp genomes and cultured isolates to detect variants absent after culture (n = 24, comprising 17 paired gastric biopsy samples and 7 paired string samples.). (ii) Gastric region-specifc SNPs, identified by comparing paired uHp genomes recovered from gastric antrum and corpus, using matched isolates as controls (n = 5). **b,** Number of unique uHp-specific SNP positions in each category, counting SNPs of all predicted effects (synonymous, non-coding and protein-changing): 21,844 in vivo-specific positions (7,550 protein-changing) and 686 gastric region-specific positions (340 protein-changing). Patients with mixed-strain infections were excluded. **c,** Circular genome map of Hp26695 (NC_018939.1, 1,667,892 bp) showing the distribution of high- and moderate-impact uHp-specific SNPs (based on snpEff^71^) across two comparison categories (**Methods**). Outer to inner tracks: (i) *in vivo*-specific SNPs, (ii) gastric region-specific SNPs. The innermost ring shows gene functional class annotations. Genes with notable SNP burden are highlighted in arc zoom boxes outside the genome ring. **d,** Gene-class enrichment analysis for SNP-bearing genes in each indicated category. Negative-binomial regression of protein-changing SNPs per gene, offset by gene length × callable comparisons; asterisks indicate Benjamini–Hochberg FDR (*<0.05, **<0.01, ***<0.001) across 24 tests; orange = higher burden, blue = lower.

We next focused on examining sequence heterogeneity that involves known virulence loci with clinical significance. Virulence-based risk stratification for gastric cancer rests on a set of Hp genes, most prominently the integrity of the *cag* pathogenicity island (cagPAI)^32^, the EPIYA motif configuration of *cagA*^4^, and the *vacA* signal / intermediate / middle-region subtype^33^ (**Fig. 3a**). uHp-seq recovered the full cagPAI and resolved *cagA* EPIYA configurations in all the 17 biopsy samples with paired isolates. While 14 of the 17 (82.4%) cases showed concordant *cagA* subtypes between those revealed by uHp and in the paired isolate genomes, patients P2c, P3a and P8c showed different *cagA* subtypes due to differences in EPIYA-C copy number (**Fig. 3b**). This discordance was driven by structural variation within *cagA* itself rather than the presence of distinct strains: a 102 bp insertion altering EPIYA-C copy number generated mixed (EPIYA-C)n (e.g. ABC/ABCC) configurations captured by uHp-seq but collapsed to a single form by culture (**Fig. 3c**). In addition, a 57 bp deletion converted an EPIYA-AB motif into an EPIYA-B configuration, producing co-existing EPIYA-B and EPIYA-AB forms in the *in vivo* sample that culture reduced to a single dominant form (**Extended Data Fig. 6a**). Although patient P2c was classified as a single-strain infection based on isolate genome, uHP-seq data shows that it harbored four distinct *cagA* subtypes (ABCC, ABCCC, BCC, and BCCC), including variants containing two or three EPIYA-C repeats (**Fig. 3d** and **Extended Data Fig. 6b**), whereas the matched cultured isolate yielded only the ABCC subtype with two EPIYA-C repeats. To validate this finding, we performed deep long-read sequencing (∼110 Gb) of native genomic DNA from P2c, without Hp enrichment or DNA amplification. This recovered three of the four cagA subtypes (ABCCC, BCCC and ABCC), in agreement with uHp-seq, and missed only the rare minor variant BCC (**Extended Data Fig. 6c,d**). Importantly, increased EPIYA-C repeat numbers are associated with elevated gastric cancer risk^4^, while emerging evidence suggests that mixed *cagA* EPIYA genotypes may also be associated with gastric cancer^34,35^, highlighting the potential clinical relevance of detecting within-host *cagA* heterogeneity. By directly resolving distinct EPIYA configurations at the single-molecule level, long-read uHp-seq reveals coexisting virulence variants that may be missed by culture-based sequencing. Two additional patterns of *cagA* heterogeneity were identified by uHp-seq. In patient W3, *cagA* was detected in only a subset of the Hp population, indicating the coexistence of *cagA*-positive and *cagA*-negative strains. In patient P12f, approximately 80% of the Hp population carried two copies of *cagA*, with the second copy being truncated (**Extended Data Fig. 6e**). Across a total of 26 *cagA*-positive biopsy samples, five had mixed *cagA* subtypes detected by uHp-seq (**Fig. 3e, Extended Data Fig. 6f**).

**Figure 3.**
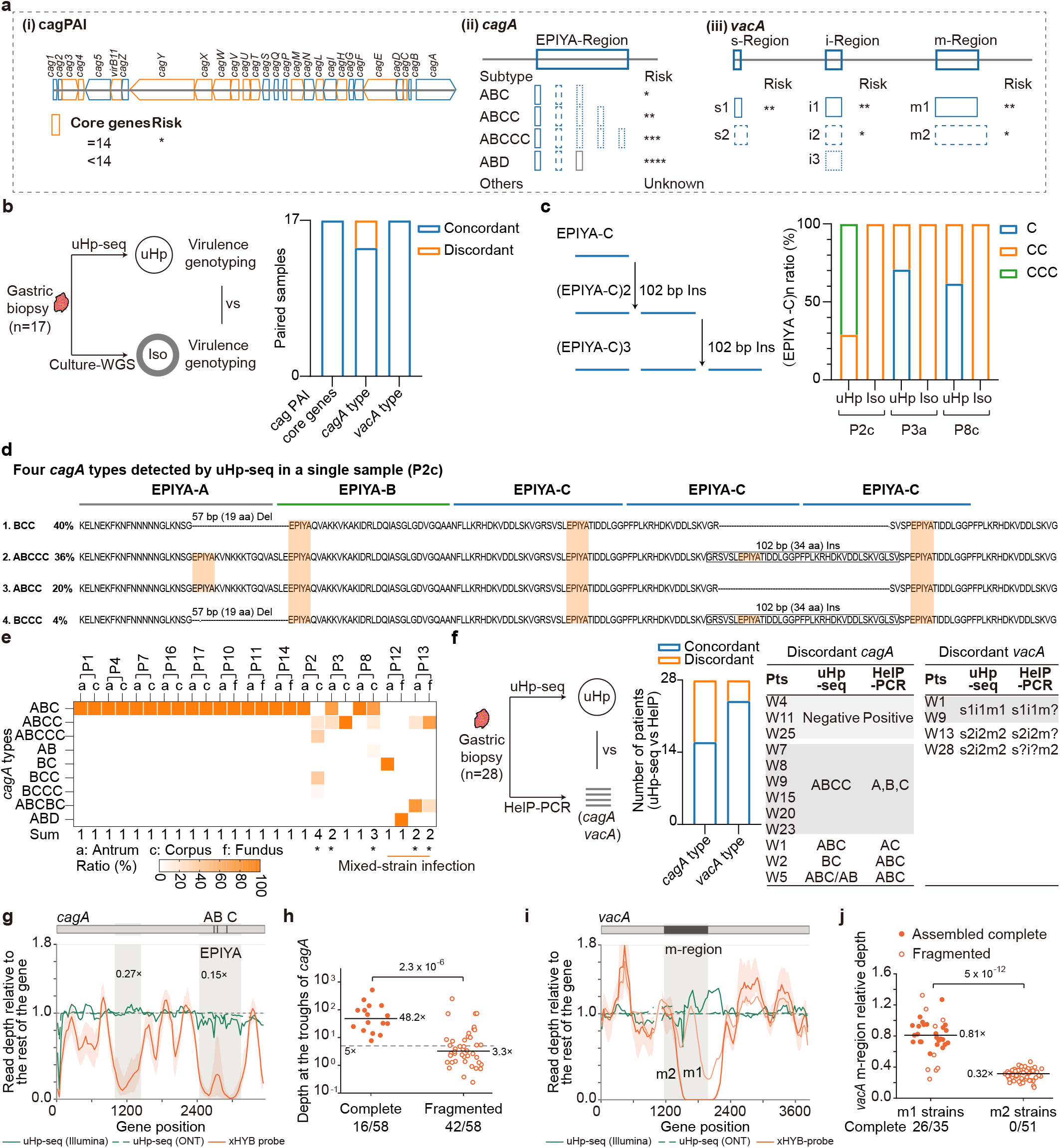
uHp-seq enables robust comprehensive virulence profiling of Hp. **a,** Schematic overview of the framework for Hp virulence profiling at gene level, integrating cagPAI integrity, *cagA* subtype, and *vacA* subtype to infer virulence potential. **b,** Comparison of virulence genotypes between uHp genomes and matched cultured isolates. Discordant *cagA* subtypes were identified in two patients. **c,** A 102 bp insertion altered the copy number of the EPIYA-C motif, generating mixed (EPIYA-C) n forms in uHp genomes that were not recovered from paired isolates. **d,** Representative example of complex *cagA* subtype heterogeneity detected by uHp-seq. Up to four distinct *cagA* subtypes were identified in the uHp genome from patient P2c. **e,** Heterogeneity of *cagA* subtype composition within samples, across gastric regions and patients. Distinct *cagA* subtypes coexisted either in single-strain infections (P2c, P3a, P8c) or in mixed-strain infections (P12 and P13). **f,** Comparison of *cagA* and *vacA* genotyping between uHp-seq and the most recently developed PCR-based clinical approach, HelP-PCR. Bars indicate the number of samples with concordant and discordant genotyping results between the two methods. The discordant genotyping results are detailed at right. **g**, Relative read depth across *cagA* obtained by uHp-seq using ONT or Illumina sequencing, or by xHYB-probe probe-based target enrichment. **h,** Read depth at the *cagA* trough regions identified in **g** in samples with complete or fragmented *cagA*. **i,** Relative read depth across *vacA* using the same sequencing and enrichment methods as in **g**. **j,** Relative read depth at the *vacA* m-region in Hp strains classified as m1 or m2.

The *in vivo* heterogeneity of *cagA* and *vacA* can also be assessed using culture-independent PCR-based methods, such as the multiplex PCR assay Deeplex HelP (HelP-PCR), a commercial assay targeting both Hp *cagA/vacA* genotyping and antimicrobial resistance (AMR) detection^21^. To evaluate this, a separate 28 biopsy samples from 28 patients were analyzed using both uHp-seq and HelP (**Fig. 3f**). Twelve samples (43%) showed discordant *cagA* genotyping results. Among these, three appear to be false-positive calls by HelP due to contamination, PCR or the reliance on short read sequencing. Compared to uHP-seq that can directly readout cagA genotypes by long sequencing reads, HelP had one sample carrying an EPIYA-ABC genotype misidentified as EPIYA-AC, and another carrying an EPIYA-BC genotype misidentified as EPIYA-ABC. Six samples harbored two EPIYA-C repeat counts that could not be correctly resolved by HelP-PCR likely because the repeat region could not be accurately resolved by the Illumina short-read sequencing used in HelP-PCR. One mixed *cagA* subtype infection was also missed by HelP-PCR because of the limited ability of short-read sequencing to resolve mixed repeat structures.

Three samples showed discordant *vacA* genotypes because HelP-PCR failed to amplify the s, i, or m region, resulting in incomplete genotyping due to primer biases, a long-standing limitation of PCR-based *vacA* typing^36^. These results demonstrate that culture-independent uHp-seq outperforms PCR-based methods for *cagA* and *vacA* genotyping by providing more accurate characterization of virulence gene diversity and heterogeneity.

We next compared uHp-seq with probe-based target enrichment using publicly available sequencing data generated with the Qiagen xHYB (xHYB-probe) Hp whole-genome capture panel^24^. Although both approaches recovered *cagA* and *vacA*, their read-depth profiles differed substantially. uHp-seq showed relatively uniform read depth across *cagA* with both ONT and Illumina sequencing, whereas xHYB showed pronounced troughs at two *cagA* loci, one of which encompassed the EPIYA region (**Fig. 3g**). Samples with coverage troughs at these two loci result in more fragmented assembly of *cagA* (**Fig. 3h**), indicating that uneven target enrichment may contribute to incomplete *cagA* recovery and preclude reliable resolution of EPIYA configurations. A similar coverage bias was observed for *vacA*: xHYB showed reduced read depth in the m-region, particularly in m2 strains, whereas uHp-seq maintained more uniform read depth across this region (**Fig. 3i**). As a result, none of the 51 m2 strains had complete *vacA* recovered by xHYB (**Fig. 3j**). These results indicate that probe-based enrichment can introduce region- and genotype-dependent coverage biases that affect the recovery and genotyping of Hp virulence genes. Beyond *cagA*, uHp-seq recovered genomes also enable assessment of other virulence genes such as *vacA*, *babA*, *sabA*, *hopQ*, and *htrA* (**Extended Data Fig. 6g**).

### uHp-seq enables full-length AMR gene profiling across six antibiotics used for Hp eradication

Selecting an effective Hp eradication regimen depends on knowing the strain’s antibiotic resistance profile^6–8^. Standard triple therapy and bismuth quadruple therapy combine a proton pump inhibitor with two of the following six antibiotics: amoxicillin (AMX), clarithromycin (CLR), metronidazole (MNZ), tetracycline (TET), levofloxacin (LEV), or rifabutin (RFB)^37^. However, rising resistance has driven first-line success rates below 80% in many regions^9^, and second- and third-line regimens require resistance information that cannot reliably be obtained without culture. For AMR-associated variants, the diagnostic gap is created by limited locus coverage of PCR as well as primer failure with highly variable resistance genes. For CLR and LEV, resistance is dominated by a small number of well-characterized mutations in 23S rRNA and *gyrA* that hotspot PCR panels cover reliably. For AMX, MNZ, TET, and RFB, however, resistance is determined by variants distributed across multiple loci, and predefined hotspot panels including the widely used commercial multiplex-PCR panels (PyloriAR^20^, HelP^21^) leave substantial resistance signal undetected. Here we evaluated whether genome-wide recovery via uHp-seq could close this gap across the full eradication antibiotic resistance genes.

First, to evaluate the impact of target coverage on AMR prediction, we compared resistance prediction accuracy between the variant set targeted by PyloriAR^20^ (a commercial test) and all AMR-associated variants recovered from complete genomes, using 96 isolates with paired whole-genome sequencing and MIC data from five previous studies^38–42^ (**Fig. 4a**; **Methods**). For CLR and LEV, both strategies performed equally well because the four canonical hotspot mutations in 23S rRNA and *gyrA* are well covered by PCR. For MNZ and TET, incorporating additional variants not covered by hotspot loci targeted by PyloriAR significantly improved sensitivity while maintaining the level of specificity (**Fig. 4b**). For AMX, all 4 resistance-associated variants across the full *pbp1A* coding sequence can be recovered from complete genomes, whereas PyloriAR’s targeted panel covers only 3 of them; the additional variants captured drove a significant improvement in sensitivity and accuracy against phenotypic MIC (**Fig. 4b**). This gain in sensitivity is accompanied by a slight decrease in specificity, reflecting the inclusion of variants with lower individual effect sizes; however, overall accuracy against phenotypic MIC is significantly higher than the targeted PCR approach (**Fig. 4b**). For RFB, both strategies performed similarly because they used the same *rpoB* resistance-associated mutations, although both showed relatively low sensitivity. Strong RFB resistance mutations are rare and were not observed in this cohort. Additional resistance-associated mutations may be identified in larger studies and can be readily incorporated into the uHp-seq analysis pipeline based on its reliable recovery of near complete Hp genomes (**Fig. 1**).

**Figure 4.**
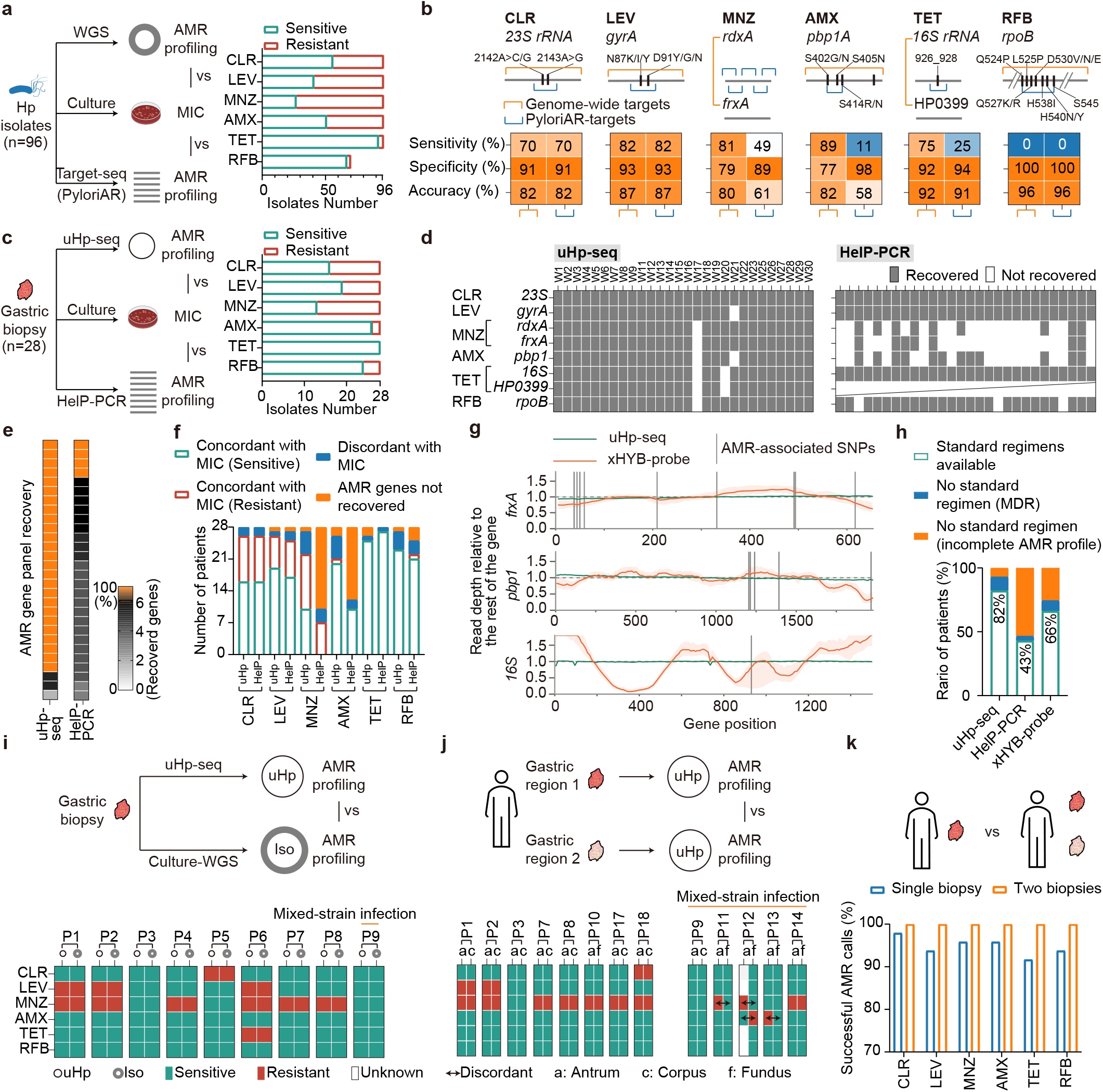
uHp-seq improves antibiotic resistance detection by recovering a broader repertoire of resistance-associated variants and genes. **a,** Public Hp datasets with matched whole-genome sequencing (WGS) and MIC measurements were used to evaluate whether expanded resistance-associated targets improve genomic AMR prediction. **b,** Comparison of antimicrobial resistance (AMR) detection performance between genome-wide WGS and commercial PCR-targeted methods PyloriAR. PyloriAR was simulated by *in silico* PCR. Genome-wide analysis showed performance comparable to PyloriAR for clarithromycin (CLR), levofloxacin (LEV), and rifabutin (RFB) resistance. Genome-wide profiling improves prediction of metronidazole (MNZ), amoxicillin (AMX) and tetracycline (TET) resistance by recovering full-length resistance genes and additional resistance-associated variants. **c**, Design for comparing AMR detection performance between genome-wide uHp-seq and commercial PCR-targeted methods HelP-PCR. **d,** AMR gene recovery by uHp-seq and HelP-PCR across the validation cohort. *HP0039* is not included in the HelP-PCR panel. **e,** Complete AMR panel recovery by uHp-seq and HelP-PCR. Complete (yellow) indicates recovery of all AMR genes; each square represents one patient. **f**, Number of patients benefiting from concordant AMR calls by uHp-seq or HelP-PCR compared with MIC. A patient was considered to benefit when the AMR call was concordant with the corresponding MIC result. More than 50% of patients had no MNZ or AMX calls by HelP-PCR because the target genes could not be successfully recovered by PCR. **h,** Relative read depth across the indicated AMR-associated genes obtained by uHp-seq in the validation cohort or by Qiagen xHYB (xHYB-probe) probe-based hybridization capture in a previously published study. Vertical lines indicate the positions of AMR-associated SNPs. **i,** Number of patients with an available standard treatment regimen or without an available regimen due to multidrug resistance (MDR) or incomplete AMR profiles, as determined by uHp-seq, HelP-PCR, or xHYB-probe. uHp-seq and HelP-PCR were evaluated using the same validation cohort, whereas xHYB-probe data were obtained from the published study described in **h**. Non-FFPE and FFPE samples were combined for xHYB-probe because no significant difference was observed between them. A minimum read depth of 5× was required for SNP calling. **i,** Comparison of AMR profiles between paired uHp genomes and cultured isolates recovered gastric corpus. **j,** Comparison of AMR profiles between paired uHp genomes recovered different gastric regions. Regional discordance in resistance profiles was observed exclusively in mixed-strain infections. **k,** Successful AMR calls when one or two biopsies per patient were analyzed. Analyzing two biopsies increased successful AMR calls across the six indicated antibiotics.

A second limitation of PCR-based commercial panels is the potential for primer-binding-site variation at resistance loci to impair amplification. Among the six major protein-coding resistance genes, four (*gyrA*, *pbp1A*, *rdxA*, *rpoB*) carry extensive sequence diversity that compromises multiplex PCR amplification across diverse strains (**Extended Data Fig. 7a**). Consistent with this, multiplex PCR assays frequently fail to recover *rdxA* and *pbp1A*, and incorporating additional targets is constrained by the fundamental complexity of multiplex primer design. In contrast, uHp-seq consistently recovered all eight major resistance loci from genomes with >70% completeness and at least seven of the eight loci from genomes with >50% completeness, covering approximately 90% of samples (**Extended Data Fig. 7b**).

To evaluate these limitations in a direct head-to-head comparison, we applied uHp-seq and HelP-PCR^21^, targeting a broader set of AMR loci than earlier panels such as PyloriAR, to an independent cohort of 28 biopsy samples from 28 patients with matched MIC measurements (**Fig. 4c**). Hp genomes were reconstructed from all 28 samples, with 26 achieving >99% genome completeness (**Extended Data Fig.7c-d**). Recovery of resistance-associated genes was comparable between uHp-seq and HelP-PCR for CLR, LEV, TET, and RFB. In contrast, uHp-seq successfully recovered the genes associated with MNZ and AMX resistance in 27 of 28 and 26 of 28 samples, respectively. For MNZ, either *rdxA* or *frxA* failed to amplify with HelP-PCR in 22 of 28 samples, while the AMX-associated resistance gene failed to amplify in 16 of 28 samples (**Fig. 4d**, **Extended Data Fig. 7e**). Consequently, uHp-seq recovered all AMR-associated genes and generated complete AMR profiles (resistance predictions for all six antibiotics) for 25 of 28 patients, compared with only 4 of 28 using HelP-PCR. (**Fig. 4e, Extended Data Fig. 7f**). We next compared the predicted AMR profiles with phenotypic MIC results. For CLR, LEV, TET, and RFB, both uHp-seq and HelP-PCR made concordant AMR calls against MIC. For MNZ and AMX, however, uHp-seq showed substantially higher concordance, primarily because the corresponding resistance genes were not successfully amplified by HelP-PCR (**Fig. 4f**). When samples with failed AMR calls were excluded, uHp-seq and HelP-PCR showed equivalent performance (**Extended Data Fig. 7g**), confirming that uHp-seq’s advantage lies in robust genome coverage and amplification reliability rather than variant calling accuracy.

To further evaluate the performance of uHp-seq in AMR detection, we compared it with the Qiagen xHYB (xHYB-probe) kit, a probe-based hybridization capture method that attempts to recover the whole Hp genome, based on the publicly available sequencing data^24^. Unlike PCR-based AST, which targets predefined resistance-associated mutations, probe-based enrichment enables broader recovery of AMR-associated genes. uHp-seq and xHYB-probe showed comparable read depth at AMR-associated SNP sites in *23S, gyrA, rdxA,* HP0399, and *rpoB* (**Extended Data Fig. 7h**). However, xHYB-probe exhibited reduced read depth at resistance-associated SNP sites in *frxA, pbp1A,* and *16S* compared with uHp-seq (**Fig. 4g**), compromising the detection of resistance-associated mutations and the completeness of AMR profiling (**Extended Data Fig. 7i**). As a result, uHp-seq would support standard treatment regimen (American College of Gastroenterology guideline-recommended Hp eradication regimen^37^) determination for 82% of patients in the validation cohort, higher than HelP-PCR (43%) and xHYB-probe (66%) (**Fig. 4h, Extended Data Fig. 7j, Methods**).

In contrast to virulence loci, where culture reshapes the *in vivo* variant landscape, AMR profiles were highly concordant between uHp-seq and paired culture-derived whole-genome sequencing in both single-strain and mixed-strain infections from the same gastric region, with no evidence that culture systematically selects for the SNPs used for determining resistance in this study (**Fig. 4i**, **Extended Data Fig. 7k,l**). AMR profiles were also largely concordant between paired uHp genomes from different gastric regions, except in some mixed-strain infections where regional differences in strain composition produced distinct resistance profiles (**Fig. 4j**), further reinforcing that within-host AMR heterogeneity reflects strain structure rather than regional genomic variation. Integrating AMR profiles across multiple gastric regions improved resistance determination for individual antibiotics (**Fig. 4k**), enabling complete resistance assignments for all 14 patients with biopsies collected from at least two gastric regions.

### uHp-seq from minimally invasive gastric string samples delivers actionable AMR profiles

To broaden the use of uHp-seq, we performed a study of gastric string samples from esophageal string test (EST), in which a swallowed capsule retrieves gastric secretions and mucus along its length^43^, representing a minimally invasive alternative to endoscopic biopsy for Hp genomics. We applied uHp-seq to 19 string samples from patients including 11 refractory Hp infection following at least two failed eradication regimens. Paired isolates, MIC data, and clinical outcome data were available for retrospective validation in 10 of 19 patients (**Fig. 5a**). Beyond reduced invasiveness, gastric strings traverse the full proximal–distal axis of the stomach, integrating Hp populations across fundus, corpus, and antrum simultaneously, potentially capturing regional heterogeneity that single or limited biopsy sampling misses.

**Figure 5.**
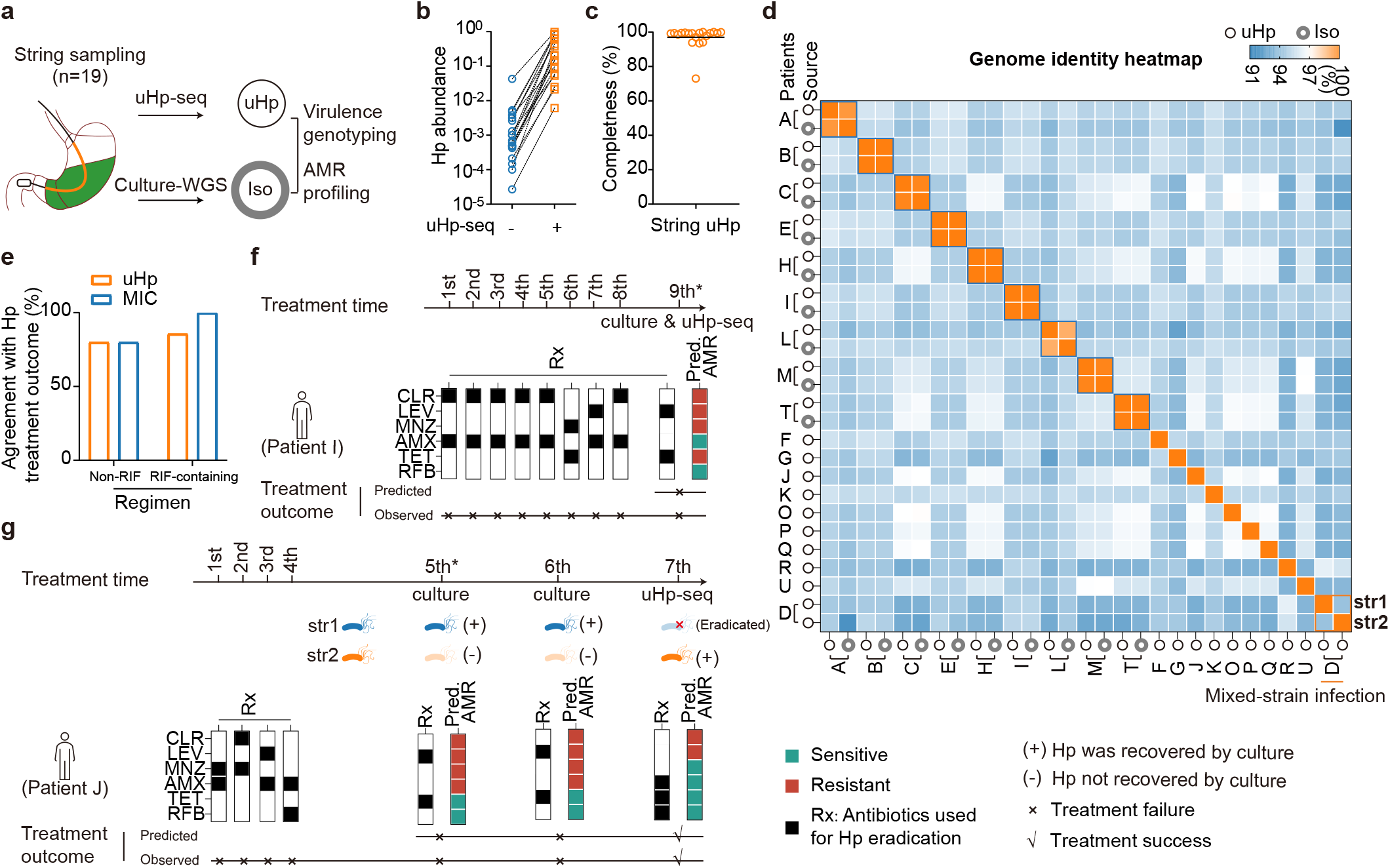
Clinical utility of uHp-seq in minimally invasive gastric string samples for genomic-guided Hp eradication therapy: retrospective clinical validation. **a,** Study design for retrospective clinical validation of uHp-seq for genomic-guided Hp treatment. **b,** uHp-seq effectively enriched Hp abundance in gastric string samples (n=19). **c,** uHp genomes recovered from string samples achieved high genome completeness (n=19). **d,** Average nucleotide identity between uHp genomes recovered from string samples and matched cultured isolates. Multiple co-existing strains were identified in patient D. **e,** Comparison of concordance between uHp-seq–predicted antibiotic resistance profiles and treatment outcomes versus concordance between phenotypic MIC measurements and treatment outcomes. Predicted uHp AMR profiles showed high agreement with treatment outcomes for non-rifabutin-containing regimens. **f,** Clinical case study of patient I. Following eight failed Hp eradication attempts performed outside the United States, uHp-seq correctly predicted resistance associated with failure of the ninth treatment regimen. **g,** Clinical case study of patient J. Gastric biopsies collected during the fifth and sixth eradication courses yielded the same cultured Hp strain (Str1). String sampling immediately after the sixth treatment followed by uHp-seq identified a second strain (Str2), confirming mixed-strain infection. The fifth and sixth regimens used the same antibiotics but different acid suppression (pantoprazole versus vonoprazan), and AMR profiles were predicted based on both strains. After the sixth course, Str1 was cleared, whereas Str2 persisted, resulting in treatment failure. The seventh regimen comprised RFB, AMX/TET, vonoprazan, and bismuth for 3 weeks; RFB was discontinued after 2 weeks and 3 days, and AMX was switched to TET after 2 weeks. Based on the remaining Str2, uHp-seq predicted treatment success, which was subsequently confirmed by a negative Hp stool antigen test.The asterisk indicates the time point of recruitment into the study. The center line in each dot plot indicates the median.

uHp-seq efficiently depleted host DNA and enriched Hp reads from all 19 string samples (**Fig. 5b**, **Extended Data Fig. 8a**), recovering high-completeness Hp genomes across all samples (**Fig. 5c**). Reconstructed string-uHp genomes showed high ANI to paired cultured isolates (**Fig. 5d**), confirming faithful genome recovery from gastric string samples. String samples also revealed mixed infections, including string sample D (**Fig. 5d**), consistent with the within-host strain heterogeneity documented in biopsy samples and further illustrating the advantage of stomach-wide sampling.

Virulence genotyping and AMR profiles derived from string sample-based uHp genomes were comprehensive and largely concordant with paired Hp isolates cultured from the same patient (**Supplementary Table 7, Extended Data Fig. 8b**). Predicted AMR profiles showed high consistency with phenotypic MIC measurements (**Extended Data Fig. 8c**). uHp-seq resistance predictions largely explained treatment failures and successes in longitudinally monitored patients, demonstrating that culture-free genomic profiling alone can provide clinically actionable information (**Fig. 5e; Methods**).

For example, in patient I, who underwent nine courses of Hp eradication therapy, the string-uHp AMR profile was consistent with the observed treatment history in the context of persistent colonization. The resistance profile explained failure of the eighth regimen by identifying resistance to CLR and further predicted failure of the ninth regimen due to persistent LEV and TET resistance, which was confirmed by clinical Hp testing (**Fig. 5f**).

In patient J, who underwent seven courses of treatment, genome comparison of string-uHp sample collected immediately after the sixth treatment with prior cultured isolates indicated mixed-strain infection, in which one strain was eradicated by the sixth treatment while the other persisted (**Fig. 5g, Extended Data Fig. 8d**). Based on the string-uHp resistance profile, the seventh regimen was predicted to succeed, which was in fact shown by the subsequent clinical Hp test outcome.

## Discussion

uHp-seq enables highly sensitive, culture-free recovery of Hp genomes directly from clinical specimens, yielding near-complete genomes from biopsies and minimally invasive string samples. By recovering Hp genomes as they exist in vivo, uHp-seq opens new opportunities to study the native biology of Hp host interaction and within-host adaptation that culture has obscured, while also enabling more precise and scalable genome-guided eradication therapy.

Decades of virulence-factor catalogs^44–49^, pangenome studies^50–53^, Hp adaptation studies^54–58^ and cancer risk associations^59–64^, including large-scale resources such as *Hp*GP^28^, have been built on cultured isolates and have therefore inherited the limitations of culture-based approaches. Beyond individual loci, within-host strain heterogeneity is substantially more complex than previously appreciated: mixed infections, regional structuring along the proximal– distal axis of the stomach, and individual informative loci such as *cagA* that harbor up to four distinct configurations within a single patient. In addition, uHp-seq identified truncating variants in multiple genes related to TonB-dependent iron and nickel acquisition that were absent from the corresponding cultured isolates. Because Hp encodes multiple TonB–ExbB–ExbD systems and FrpB/FecA-family transporters with potential functional redundancy^65^, disruption of individual components may attenuate rather than abolish metal acquisition, potentially reflecting adaptation to the nutrient landscape of the gastric niche. Their disappearance after culture further suggests that such *in vivo-*adapted variants may be selectively disadvantaged under laboratory growth conditions, illustrating how culture can erase biologically relevant within-host variation before genomic analysis. These findings argue that core questions of Hp population structure, host adaptation, and within-host evolution should be revisited with culture-free genomic approaches.

For virulence genotyping, uHp-seq has important advantages over existing approaches: it captures the *in vivo* virulence landscape that culture distorts, and it resolves full locus configurations including the intact ∼40 kb cagPAI, structural variants within *cagA*, and the highly variable adhesin repertoire, which PCR panels do not reliably cover. In addition, given that EPIYA-C copy number is associated with CagA activity^66^, uHp-seq reliably resolved EPIYA-C copy-number variation that was missed by probe based target-enrichment due to hybridization biases. The most striking virulence finding is that individual hosts simultaneously harbor multiple co-existing *cagA* EPIYA variants with distinct oncogenic potential, a within-host heterogeneity that culture collapses to a single dominant type. EPIYA configuration is a primary determinant of CagA-mediated oncogenic signaling through SHP-2^67^, and the co-existence of variants with distinct oncogenic potential within a single host means that culture-based EPIYA typing misrepresents the true virulence landscape of the infection. A patient presumed by a clinician to carry a low-risk EPIYA configuration may simultaneously harbor a high-risk variant that was not recovered by culture. Current gastric cancer risk models built on culture-derived EPIYA classification may therefore be systematically underestimating risk in patients whose most oncogenic variant is culture-disadvantaged. uHp-seq provides the genomic substrate to build risk scores that reflect the full *in vivo* virulence distribution rather than its culture-selected proxy^61,68^.

The culture-biased view of Hp virulence extends beyond *cagA*. Whole-genome recovery across all virulence loci reveals that the adhesin repertoire (including *babA*, *sabA*, *hopQ*, and *htrA*) has been similarly affected. In addition, we found that genome-wide strain typing similarly exposes limitations of current practice: PCR-based 7-gene MLST^69^, the standard culture-free typing approach, recovered all seven housekeeping genes (*atpA*, *efp*, *mutY*, *ppa*, *trpC*, *ureI*, and *yphC*) in only 22% of samples using HelP-PCR, compared with 93% using uHp-seq. While HelP-PCR provided only coarse geographic population assignments, uHp-seq enabled high-resolution genome-wide strain typing into 17 subpopulations as defined by the *Hp*GP^28^. Using the high-resolution geographic population classification system, MLST achieved only 42–50% concordance with genome-wide geographic population structure analysis across the two cohorts. (**Extended Data Fig. 9**), with implications for the population-level inferences about Hp diversity and cancer risk associations that MLST-based studies have generated. Taken together, these findings indicate that the full virulence landscape of Hp, across cagPAI, EPIYA configuration, adhesins, and strain identity, has been assessed through a culture-distorted lens, and that uHp-seq provides the substrate to revisit it.

For AMR-associated variants, our study showed that the diagnostic gap here owes to the restricted locus coverage of commercial PCR and probe panels, as well as primer failure on highly variable resistance genes. uHp-seq genome-wide recovery outperforms targeted PCR panels for AMX and MNZ specifically, where resistance is determined by variants distributed across full gene sequences rather than a small number of canonical hotspots. Reliable determination of AMX susceptibility is increasingly important with the introduction of vonoprazan–amoxicillin dual therapy as a first-line treatment option^37,70^. This regimen uses a single antibiotic, reduces exposure to other antibiotics, and is generally well tolerated. Because its efficacy relies directly on AMX activity, accurate detection of AMX resistance before treatment could help guide appropriate use and avoid treatment failure. For CLR and LEV, where resistance is dominated by a handful of well-characterized mutations, targeted panels perform equivalently, confirming that whole-genome recovery adds value precisely where PCR’s structural constraints have the largest effects. Another advantage of uHp-seq over multiplex PCR panels is that newly emerging resistance variants are readily incorporated into profiling as the genome is recovered whole, without primer redesign. In contrast, multiplex PCR panels must be deliberately redesigned to add new targets, a slow process constrained by primer complexity in variable regions like *pbp1A* and *rdxA*.

Beyond accurate resistance genotyping, the gastric string cohort reveals that string-based uHp-seq is not merely a less invasive alternative to endoscopic biopsy but a more comprehensive one. By traversing the full proximal–distal axis of the stomach, the string integrates Hp populations across fundus, corpus, and antrum simultaneously. The regional virulence and AMR heterogeneity documented in biopsy samples, where distinct *cagA* subtype compositions and resistance profiles were identified across gastric regions in patients with mixed-strain infections (∼10% of samples), highlights that single-site sampling is inherently incomplete for characterizing the true within-host Hp landscape. Multi-region biopsy sampling generated complete AMR profiles for all 14 patients versus single-site sampling, and the gastric string achieves the equivalent of that multi-region integration in a single procedure not requiring endoscopy. For both virulence risk stratification and resistance-guided eradication therapy, stomach-wide sampling is clinically advantageous, and the gastric string is currently the only practical way to achieve it without endoscopy.

In this study, uHp-seq requires Hp at ≥10⁻ relative abundance to reconstruct Hp genomes, a threshold met by ∼90% biopsies and string samples. While we demonstrated uHp-seq on gastric biopsies and string samples, extension to other clinically relevant sample types, including fixed gastric biopsies, gastric juice, stool, and saliva, will be optimized in future work. Although the gastric string cohort provides informative retrospective validation of uHp-seq-guided therapy selection, prospective validation studies in independent cohorts with integrated treatment-outcome data will be required to establish the clinical utility of routine deployment of uHp-seq.

Despite these constraints, uHp-seq establishes a foundation for culture-free Hp genomics, making the *in vivo* within-host bacterial population accessible at clinical scale. By recovering genomes directly from host-dominated specimens, this approach opens a practical path toward precision eradication therapy, more accurate gastric cancer risk stratification, and potentially a broader framework for direct genome recovery from other clinical specimens where culture has long been the only option.

## Methods

### Sample collection

Gastric biopsy and EST samples were obtained from multiple cohorts and studies. All samples were collected with informed consent and under the respective institutional review board (IRB) approvals, unless otherwise specified below. Collection 1, 2: 41 gastric biopsies were collected at the University of Magdeburg (IRB no. 80/11)^72^ and Collection 3:18 gastric biopsies were collected at the Veterans Affairs Medical Center in Nashville (IRB no. 5910)^73^, during routine endoscopy. Collection 4: 6 gastric biopsies acquired from National Cancer Institute were from the *Hp*GP, which were collected at the A. C. Camargo Cancer Center, São Paulo, SP, Brazil (Protocol no. 2134/15)^74^. 4 gastric biopsies were collected at the Pontificia Universidad Católica de Chile (ClinicalTrials no.NCT03188406). Collection 5: 28 remnant gastric biopsy specimens submitted to the Montefiore Medical Center Clinical Microbiology Laboratory for Hp culture and antimicrobial susceptibility testing were used for research (IRB no. 2018-9702), which waived the requirement for informed consent owing to the use of remnant clinical specimens. 24 gastric tissue samples were collected at Mount Sinai from individuals undergoing gastric bypass surgery. The study was exempted from IRB evaluation. 19 EST samples were collected at the National Institute of Allergy and Infectious Diseases as part of a study evaluating the EST for Hp detection and antimicrobial resistance characterization (IRB no. 002302; ClinicalTrials no. NCT06871579). All samples were stored at −80 °C until processing. Patient demographics and clinical metadata are summarized in **Supplementary Table 1-2**. No personal identifiable information was used in the scope of this work.

### Esophageal string test (EST) sample collection

The EST is a minimally invasive, FDA-registered device (EnteroTracker; EnteroTrack) consisting of an adsorbent nylon fiber string housed within a weighted gelatin capsule, designed to collect biofluid from the upper gastrointestinal tract without endoscopy^75^. The 140-cm string was used for all samples, and the string was left in place for 1 hour prior to retrieval. Patient height was used to estimate the distance to the lower esophageal sphincter (LES) using the manufacturer-provided formula (6.7 + 0.226 x height [cm]). The portion of string distal to this point was used for analysis.

### Hp isolation from gastric specimens and culture

**Hp isolation at Vanderbilt University:** Hp was isolated from antral and corpus biopsies, as previously reported^76^. Briefly, specimens were homogenized in sterile saline. Serial dilutions were plated onto selective Trypticase Soy Agar (TSA) with 5% sheep blood. Agar plates were incubated under microaerobic conditions at 37°C for 4 days. Hp was confirmed by positive oxidase and urease tests and typical appearance on a Gram stain. **Hp isolation at Montefiore Medical Center:** Hp culture was performed for routine clinical care at the Montefiore Medical Center Clinical Microbiology Laboratory, in accordance with standard operating procedures. Gastric biopsy specimens were plated onto Columbia with nalidixic acid and 5% sheep blood agar plates (BD BBL, Sparks, Maryland) and incubated under microaerophilic conditions at 37°C for up to 7 days. Identification of Hp was confirmed by positive catalase, oxidase, and rapid urease testing. **Hp isolation at National Institute of Allergy and Infectious Diseases:** After EST, gastric antral biopsies from patients with confirmed Hp infection were cultured on blood agar (BHI + 5% sheep blood) and on a selective CVA (cefsulodin-vancomycin-amphotericin B) supplemented plate, both incubated under microaerophilic conditions at 35°C. Hp and *H. bilis* reference strains were run in parallel on BHI plates as quality controls. Colonies with morphology consistent with Hp were confirmed by MALDI-TOF mass spectrometry.

Hp isolates stock were recovered on TSA with 5% sheep blood at 37°C in 5% CO_2_ for 2-3 days, all colonies were collected into one tube for DNA extraction.

### DNA extraction

**Hp isolates:** Collect the colonies from TSA blood plates, and DNA was extracted using the DNeasy Blood & Tissue Kit (Qiagen, Cat # 69506), QIAamp DNA Mini kit (Qiagen, Cat # 51306) or MasterPure^TM^ Complete DNA and RNA Purification Kit (Lucigen, cat. no. MC85200) following the manufacturer’s protocol. **Gastric biopsy:** 1/3 of each regular biopsy was minced and processed for total DNA extraction using the DNeasy Blood & Tissue Kit (Qiagen, Cat # 69506) or PureLink Genomic DNA Kit (Invitrogen Inc. Cat # K182001) following the manufacturer’s tissue protocol. DNA was eluted in low-EDTA buffer or nuclease-free water. **Fecal DNA:** extracted with PowerFecal Pro DNA Kit following it protocol (Qiagen# 51804). **String samples:** Total DNA was extracted using the MasterPure^TM^ Complete DNA and RNA Purification Kit (Lucigen, cat. no. MC85200) according to manufacturer’s instruction for cell samples. Briefly, the gastric segment of the string was placed in 600µL of Tissue and Cell Lysis Solution with 2 µL of Proteinase K and vortexed briefly, after which a sterile slide was used to strip adherent fluid from the string surface. The final DNA pellet was resuspended in 40 µL of TE buffer.

### DNA mocks

Human DNA (extracted from human lymphoblastoid cell line culture) and DNA extracted from Hp-negative human stool (confirmed by both the urea breath test and stool antigen test) were used separately as background DNA. Human DNA, Hp-negative stool DNA, and Hp26695 DNA were separately sheared to ∼10 kb using g-TUBEs (Covaris, #520079). Mock mixtures were prepared by spiking Hp26695 DNA into either human DNA or Hp-negative stool DNA at Hp DNA abundances ranging from 1 to 1.0 × 10⁻, with a total DNA input of 1 µg per mixture.

### Restriction enzyme digestion and DNA fragment analysis

Human, Hp, or mock DNA (200 ng – 3ug) was incubated with methylation-sensitive restriction endonucleases HhaI, NlaIII, and MboI alone or combination as indicated in corresponding figures (New England Biolabs, Cat # R0139S, Cat # R0125, Cat # R0147M) at 37 °C for 30 min. The REase was then heat-inactivated at 65 °C for 20 min, and DNA fragment size distributions were analyzed using a TapeStation or 0.9% agarose gel electrophoresis.

### QC of input DNA

Input DNA was required to be high quality, pure total genomic DNA meeting the following criteria: (a) purity measured by Nanodrop, OD 260/280 of 1.8 and OD 260/230 of 2.0–2.2; (b) average fragment size measured by electrophoresis (>10 kb); (c) Input mass measured by Qubit, ideally 200 ng in total.

### qPCR measure Hp abundance in input DNA

A standard curve was generated using human and Hp DNA mock samples. Mock DNA or Input DNA was diluted to 10 ng/µl for qPCR. Each 10 µl qPCR reaction contained 5 µl TaqMan Fast Advanced Master Mix (Applied Biosystems, Cat # 4444556), 0.25 µl forward primer (OYJL234) (5’-GAGCTGTCTCAACCAGAGATT-3’), 0.25 µl reverse primer (OYJL235) (5’-GCAGTGCTAAGTTGTAGTAAAGGTC-3’), 0.125 µl probe (OYJL236) (5’-FAM/TG GAG GTG AAA TTC CTC CTA CCC G/3’-TAMRA) ^77^, 10 ng template DNA and nuclease-free water. Reactions were run on a QuantStudio 6 Flex using the fast cycling program: 50 °C for 2 min; 95 °C for 20 s; followed by 40 cycles of 95 °C for 1 s and 60 °C for 20 s.

### uHp-seq enrichment and library preparation

Genomic DNA (200 ng per sample) was co-digested with the methylation-sensitive restriction endonucleases HhaI, NlaIII, and MboI (New England Biolabs, Cat # R0139S, Cat # R0125, Cat # R0147M) for 90 min while fragmenting unmethylated background DNA. Then, 1× AMPure XP (AXP) beads were used to exchange the cutting buffer into TE buffer. Long DNA fragments were then isolated on a BluePippin (Sage Science, protocol-0.75% DF Marker S1 high pass 6-10 kb vs3) using a size window of 3 to 50 kb optimized to recover long fragment Hp DNA. For long-read sequencing, recovered DNA was barcoded using 1µl MuA transposase (Oxford Nanopore Technology (ONT) Rapid Barcoding kit, RPB.114.24) prior to Nanopore sequencing on MinION or PromethION flow cells (ONT). For short-read sequencing, libraries were prepared either by Tn5-mediated tagmentation directly following size selection or by amplification from the long-read barcoded product and sequenced on Illumina platforms NovaSeq 6000.

### Microbial-enrichment method (MEM)

MEM was performed using 100 mg of fresh gastric tissue as previously described^30^. Following bead beating, the homogenized tissue was divided into two 150µl aliquots. The first aliquot was subjected to the subsequent MEM enrichment steps. The second aliquot was processed using the QIAamp DNA Microbiome Kit (Qiagen, Cat # 51704), in which human cells remaining intact in the liquid fraction after bead beating were further selectively lysed with saponin, and the released human DNA was digested with Benzonase before bacterial cell lysis and DNA extraction.

### Direct deep long-read sequencing

DNA from gastric biopsy or string samples was extracted as described above, without any host DNA depletion or enrichment procedures. ONT sequencing libraries were prepared using the Native Barcoding Kit (SQK-NBD114.24) according to the manufacturer’s protocol. Libraries were sequenced on the PromethION platform to generate sufficient sequencing depth to obtain adequate Hp reads for whole-genome assembly.

### ONT sequencing data processing

Raw ONT reads were base called and demultiplexed using Dorado (v0.4.1)^78^ with the dna_r10.4.1_e8.2_400bps_sup.cfg model for R10.4 flow cells and the emit-fast flag to produce per-barcode FASTQ files. Barcodes yielding fewer than 10 Mb of sequence were excluded. Sequencing statistics (total reads, read length N50, mean length) were computed using seqkit^79^.

### Metagenomic classification and community profiling

Demultiplexed reads were classified using Kraken2 (v2.0.9b)^80^ against the PlusPF database (comprising RefSeq archaea, bacteria, viruses, plasmids, fungi, protozoa, and the human genome). Taxonomic reports were filtered to species- and unclassified-level entries. To obtain base-resolved community profiles, Kraken2 raw output was parsed to extract per-read base-pair assignments; NCBI taxonomic lineage was appended using TaxonKit^81^ with the NCBI taxonomy database (downloaded February 2023). Community profiles were visualized as stacked bar charts in R (v4.2.0) using the tidyverse and Polychrome packages.

### uHp-seq enrichment analysis

Enrichment efficiency was quantified by comparing paired input and uHP-seq-enriched libraries for each sample using two orthogonal metrics: (1) Hp fractional abundance derived from Kraken2 classification reports^80^, and (2) Hp read mapping rate determined by aligning reads to the Hp26695 reference genome (GCF_000307795.1; NC_018939.1) with Minimap2 (v2.24; -ax map-ont)^82^, with mapping statistics extracted by SAMtools (v1.15.1) flagstat^83^. The enrichment fold was calculated as the ratio of the respective metric in the enriched library to that in the matched input library. Genome breadth of coverage and mean sequencing depth were computed with SAMtools coverage^83^.

### Hp read extraction and *de novo* genome assembly

Reads classified as Hp by Kraken2 were extracted per barcode to produce HP-only FASTQ files. *De novo* assembly was performed with Flye (v2.9)^84^ using the nano-hq flag and 30 threads. For barcodes with evidence of mixed or heterogeneous Hp populations, assembly was additionally performed in metagenome mode (meta). Assembly completeness and contamination were assessed with CheckM (lineage_wf workflow, 8 threads)^85^. Read-based genome coverage was calculated as the fraction of the matched isolate genome (or the closest publicly available reference genome when a matched isolate was unavailable) covered by at least one read, using read depths in 100-bp bins.

### Strain-level assembly refinement and ANI estimation

For each species-level haplotype-phased assembly, contigs were further resolved into strain-level assemblies using Strainberry v1.1^86^, which separates strain-specific haplotypes using long-read-supported phasing information. The resulting strain-resolved contigs were binned using the binning module of metaWRAP v1.3.2^87^, and the quality of reconstructed long-read metagenome-assembled genomes (MAGs) was assessed and refined using the bin_refinement module of metaWRAP. Pairwise average nucleotide identity (ANI) among Hp strain-level assemblies was calculated using fastANI v1.32^88^.

### Nucleotide diversity (*pi* / π) of uHp genomes and cultured isolates

Within-sample nucleotide diversity *pi* was compared between uHp genomes and cultured isolates using Illumina short-read data. Reads were deduplicated, screened against phiX, adapter-trimmed, quality-trimmed and length-filtered (minimum 75 bp), then aligned to the Hp26695 reference genome (GCF_000307795.1, NC_018939.1; 1,667,892 bp) with Minimap2 v2.30 (-a -x sr --secondary=yes) and retained at mapping quality ≥ 30 (SAMtools v1.23). To make samples directly comparable, each alignment was randomly downsampled to a common depth of 1,375,000 mapped reads with Picard DownsampleSam v3.4.0 (STRATEGY=Chained, ACCURACY=1e-4), and the analysis was repeated over 10 independent downsampling rounds. At each reference position, *pi* was calculated from the per-base read counts as the proportion of pairwise differences among the reads covering that position. Genome-wide *pi* was the mean across all reference positions, coding *pi* the mean across coding positions, and per-gene *pi* the mean across the positions of each of the 1,485 annotated genes; reported values are means across the 10 rounds.

### SNP calling and annotation

**Illumina data.** Paired-end reads were quality-trimmed with fastp (v0.23.2)^89^ and aligned to the Hp26695 reference genome (NC_018939.1) using BWA-MEM (v0.7.17)^90^. BAM files were downsampled to a target depth of 200× with SAMtools^83^. Variants were called with LoFreq (v2.1.5)^91^ at a minimum allele frequency (AF) of 0.02 and minimum site depth of 20×. **ONT data.** ONT reads were aligned to the Hp26695 reference genome (GCF_000307795.1; NC_018939.1) with Minimap2 v2.24 (-ax map-ont), downsampled to 200x mean depth, called with Clair3 v1.0.10 (model r1041_e82_400bps_sup_v420; retained at QUAL >= 20, DP >= 20, AF >= 0.05) and annotated with snpEff against a custom database (Hpylori_custom).

Because the QUAL filter discards genuine calls in lower-coverage isolates, category membership was decided from the reads rather than the filtered VCFs: candidate alleles (the panel-wide union of biallelic SNVs in the raw Clair3 VCFs^92^) were genotyped in all samples with samtools mpileup (-q 20 -Q 10), and all categories are defined on exact alleles. *In vivo*-specific SNPs required biopsy DP >= 20, AF >= 0.10, at least 5 ALT reads on both strands and deletion fraction < 0.15, with the matched isolate covered (DP >= 20) at AF < 0.05 and delta AF >= 0.10; intra-strain SNPs are the subset with biopsy AF between 0.30 and 0.70. Gastric region-specific SNPs were called in the subjects with antrum and corpus biopsies plus matched isolates, requiring one biopsy site positive (AF >= 0.20, same read support), the other covered and negative (AF < 0.20), and no covered isolate carrying the allele. Samples with multi-strain co-infection were excluded.

Gene-class SNP burden was modelled per category by negative binomial regression of protein-changing (snpEff HIGH or MODERATE^71^) events per gene on gene-class membership, with log(gene length in kb x callable comparisons) as offset; each of the eight classes was tested against all other callable genes and the resulting 24 P values were adjusted by Benjamini-Hochberg. Gene-level burden and chromosomal distribution were drawn with the R package circlize^93^.

### Geographic clade and subpopulation assignment

Geographic subpopulation assignment of Hp strains was performed using two complementary approaches. **DAPC on core-genome SNPs.** Each query assembly was aligned to the Hp26695 reference with snippy (ctgs), and a core-genome SNP alignment (632,909 informative positions) was generated with snippy-core together with 1,199 geographically annotated genomes from the *Hp*GP^28^. Queries were projected into the discriminant space of a DAPC model pre-trained on the reference panel (200 principal components, 17-class LDA) and assigned to the nearest population centroid by Euclidean distance. Assignment confidence was quantified as the relative margin (d2 − d1)/d2, where d1 and d2 are the distances to the top- and second-ranked centroids; margins >0.5 were classified as high confidence, 0.2–0.5 moderate, and <0.2 low. Subpopulation labels (17 subpopulations) were taken from the fineSTRUCTURE^94^-based classification reported by Thorell et al^28^ **MLST-based approach.** For assembled genomes, 7-gene MLST was performed using the standard Hp scheme (pubMLST Helicobacter scheme 1^95^; loci: *atpA, efp, mutY, ppa, trpC, ureI, yphC*). Each assembly was queried against the pubMLST allele database by BLASTn (≥85% nucleotide identity, ≥90% allele-length coverage); the best-matching allele per locus was selected by identity then alignment length, and the resulting profile was matched to the pubMLST sequence type (ST) table. Because STs are not themselves informative of geographic ancestry, subpopulation was inferred by allele sharing against the *Hp*GP reference panel of 1,200 fineSTRUCTURE-labelled genomes, typed by the identical BLASTn procedure: allelic proximity to each reference strain was defined as 1 − (fraction of commonly typed loci with different alleles), proximity was averaged within each of the 17 subpopulations, and the subpopulation with the highest mean proximity was reported with a confidence margin (difference in mean proximity between the top and runner-up subpopulation).

### Virulence factor analysis

Virulence genes were identified from the Prokka-annotated assemblies by BLASTp (BLAST+ 2.16.0) of all predicted proteins against a reference panel of *cagA* (WP_267286128.1), VacA (AAD07935.1) and HtrA (AAD08063.1), with E-value < 1×10⁻ ; hits were retained at ≥70% amino-acid identity, alignment length ≥300 aa and bit score >500, and the corresponding gene and protein sequences extracted with seqkit. Because assembly and annotation resolve only the dominant strain in a sample, all cag PAI and *cagA* presence/absence calls were made from reads rather than from assemblies. Raw reads were mapped with minimap2 (-ax map-ont) to the Hp26695 reference chromosome and per-base depth and breadth computed with samtools across the cag PAI (*cag1–cagA*, ∼547–583 kb) and across *cagA*, using the immediate flanking regions as a within-sample full-depth control. Samples were called negative only when depth was 0× across the entire island while flanks retained full depth. *cagA* subtype (EPIYA) and *vacA* subtype (s1/s2, m1/m2, i1/i2/i3) was determined from assembled gene sequences in two ways:(1) alignment to established reference alleles. (2) check the feature specific sequences.

### Antimicrobial resistance prediction

AMR-associated variants were detected at a curated panel of resistance loci targeting clarithromycin (CLR; 23S rRNA, both chromosomal copies), levofloxacin (LEV; *gyrA*), amoxicillin (AMX; pbp1), metronidazole (MNZ; *rdxA*, *frxA)*, tetracycline (TET; 16S rRNA, both chromosomal copies, HP0399) and rifampin (RIF; *rpoB*) (Rifabutin is used for Hp treatment and rifampin is used for MIC test). Detection of any mutation in the curated resistance mutation panel was considered indicative of resistance to the corresponding antibiotic. Resistance positions were encoded in a curated BED file (see **Supplementary table 8**) distinguishing Tier-1 positions (allele-specific, e.g., 23S rRNA A2143G/A2144G) from Tier-2 positions (codon-level, requiring amino acid annotation to confirm pathogenicity, e.g., *gyrA*, *rpoB*). For ONT raw reads, reads were aligned to the Hp26695 reference with Minimap2^82^ and variants at AMR loci called by BCFtools (v1.22) mpileup and call^83^ in haploid mode (-m), with minimum mapping quality Q20, minimum base quality Q20, and a maximum pileup depth of 100,000. Sites with depth < 20× were reported as indeterminate (ND) rather than wild-type. For Illumina data, BWA-MEM^90^ alignment and LoFreq^91^ calling were applied as described above. For genome assemblies, nucmer-based^96^ whole-genome alignment to the reference was used. All calls were annotated with snpEff^71^ and cross-referenced against the curated resistance amino acid table to resolve Tier-2 positions as confirmed resistant (R) or background variant. For CLR, 23S rRNA heteroplasmy was assessed by computing allele frequencies across both rRNA gene copies; positions with AF between 10% and 80% were flagged as heteroplasmic (R_HET), consistent with a single-copy mutation appearing at ∼50% AF in haploid-equivalent ONT pileups.

### Hp AMR detection with HelP-PCR

From the total DNA extracted, targeted amplification of Hp genomic content was performed using GenoScreen’s Deeplex HelP kit as per manufacturer’s instructions. The PCR amplicons were purified using AMPure XP magnetic beads (Beckman Coulter, USA) and quantified using the Qubit double-stranded DNA Broad Range Assay Kit (Invitrogen Inc.). Following the amplicon purification, library preparation was performed using the Nextera XT Library Preparation Kit (Illumina, San Diego, CA) following the manufacturer’s protocol with the following modifications – Hp DNA amplicons were diluted to 0.2 ng/uL in molecular grade nuclease-free water (Sigma-Aldrich, USA), for a 1 ng of DNA mass input into the library preparation workflow. AXP magnetic beads (Beckman Coulter, USA) was used to purify the Hp DNA libraries following library amplification. Final DNA library quantification was performed using the Qubit double-stranded High Sensitivity Assay Kit (Invitrogen Inc.). The libraries were then normalized to a final concentration of 2 nM each and pooled. Sequencing of these libraries was performed on the Illumina MiSeq Platform using the MiSeq v2 Micro Sequencing Kit (300 cycles; 2 x 150 bp) with 1% bacteriophage phiX control (Illumina, San Diego, CA). Sequencing FastQ files were uploaded to GenoScreen’s secure server for analysis.

### Comparison with probe-based capture enrichment xHYB-probe

To benchmark uHp-seq against hybridization-capture enrichment, we reanalyzed the public QIAseq xHYB *Hp* whole-genome sequencing dataset (ENA PRJEB110105)^24^, comprising 104 Illumina NextSeq 500 paired-end (2 × 151 bp) libraries prepared directly from 28 fresh and 70 formalin-fixed paraffin-embedded (FFPE) gastric biopsies. The deposited reads had been filtered to only preserve *Helicobacter* reads, so enrichment efficiency could not be compared; the comparison was restricted to resistance genotyping and virulence-gene recovery. The uHp-seq comparator comprised the 87 biopsy genomes in **Supplementary Table 2** that were sequenced by uHp-seq and yielded an assembly.

#### Read processing and assembly

Reads were trimmed with fastp (v0.23.2)^89^, aligned to Hp26695 (NC_018939.1) with BWA-MEM (v0.7.17)^90^., and PCR duplicates were removed with SAMtools (v1.16)^83^. All 104 libraries were assembled without depth filtering, from all non-duplicate read pairs (downsampled to 150× where deeper), with SPAdes (v3.13.1; --careful). Contigs <500 bp, and contigs assigned by Kraken2 to lineages outside *Helicobacter*, were removed from xHYB and uHp-seq assemblies alike. Libraries that failed to assemble were kept in all denominators.

#### Resistance variants

AMR-associated variants were called from xHYB reads with the uHp-seq Illumina pipeline (Antimicrobial resistance prediction), with sites below 20× reported as indeterminate; a 5× threshold was also tested as a sensitivity analysis.

#### Virulence genes

Virulence genes were scored as described for uHp-seq (Virulence factor analysis). Read depth along *cagA* and *vacA* was profiled after assigning reads to their closest RefSeq allele with DIAMOND blastx (v2.2.0), using uHp-seq libraries as no-capture controls.

### Hp MIC test

#### Gastric biopsy paired isolates

Hp susceptibility testing was performed at the Montefiore Medical Center Clinical Microbiology Laboratory, in accordance with standard operating procedures and as described previously^40^. In brief, testing was performed using gradient diffusion strips (bioMérieux, France). **String sample paired isolates**: Confirmed isolates were submitted to Mayo Clinic Laboratories for antimicrobial susceptibility testing by broth microdilution (Test ID: HELIS). Literature supported breakpoint was used for RIF susceptibility interpretation (Isolates classified as resistant at MICs >4 µg/ml)^97^. For research purposes only, EUCAST 2026 (version 16.1) breakpoints were used for interpretations for other antibiotics.

### Selection of Hp eradication regimens

Five Hp eradication regimens were considered based on the 2024 American College of Gastroenterology clinical guideline: optimized bismuth quadruple therapy (BQT; PPI, bismuth, MNZ, and TET), rifabutin triple therapy (omeprazole, RFB, and AMX), PCAB dual therapy (vonoprazan and AMX), PCAB triple therapy (vonoprazan, CLR, and AMX), and levofloxacin triple therapy (PPI, LEV, and AMX). Because each of these regimens contains either AMX or MNZ, resistance information for both antibiotics was required to comprehensively evaluate regimen availability. A standard treatment regimen was considered available if all antibiotics included in the regimen were predicted to be susceptible. Samples for which no standard regimen was identified were further classified as having no available standard regimen due to multidrug resistance or an incomplete AMR profile.

### String-sample validation cohort and treatment-outcome analysis

String samples were obtained from treatment-naive patients or refractory Hp patients following at least two failed eradication regimens, with clinical outcome data available. Treatment outcome was defined by stool antigen test. The biopsies were also collected for Hp isolation and phenotypic MIC testing. uHp-seq predicted resistance profiles were compared with phenotypic MIC data. For non-RFB-containing regimens, a regimen was predicted to fail if resistance was detected to any antibiotic included in the regimen. For RFB-containing regimens, failure was predicted only when resistance to RFB or both RFB and at least one other antibiotic in the regimen was detected. Predicted treatment outcomes were then compared with the clinical eradication outcomes.

### Statistics and reproducibility

Sample sizes were determined by specimen availability across the contributing sites. All genomic comparisons were paired within patients (biopsy versus matched isolate, or antrum versus corpus), so no randomization or blinding was applicable. Samples with evidence of multi-strain co-infection were excluded from the SNP category analyses as listed above; no other data were excluded.

Statistical tests, sample numbers and the meaning of any centre line or error bar are stated in the corresponding figure legends. Where a family of related comparisons was made, P values were adjusted by the Benjamini-Hochberg procedure and asterisks denote FDR < 0.05 (*), < 0.01 (**) and < 0.001 (***). Concordance between genomic predictions and phenotypic or clinical outcomes is reported as counts and percentages of patients. Analyses were performed in R and Python, with versions given in the corresponding Methods sections.

## Data availability

All sequencing data generated for this study have been submitted to the Sequence Read Archive under BioProject (PRJNA1511313), which will be publicly released upon publication. The hybrid-capture sequencing data were publicly available from the European Nucleotide Archive under accession PRJEB110105.

## Acknowledgement

We thank our colleagues Lei Cao, Magdalena Księżarek and Yangmei Li at the Icahn School of Medicine at Mount Sinai, for their valuable suggestions on this study. We also thank P. Malfertheiner at Otto-von-Guericke Universität Magdeburg, Giulio Quarta Hackensack University and Zhan Gao at Rutgers University, Thenuka Jayatilaka at National Institutes of Health for helping collect gastric specimens. Additionally, we thank Jonas Sean Adrian Lopez, Sheena Shiwlani, and Adonis Mateoii for their efforts in the acquisition and evaluation of gastric tissue that was used in these studies. This work was primarily supported by a seed fund (G.F.) from the Icahn School of Medicine at Mount Sinai. The bacterial epigenome analyses were supported by grant no. R35 GM139655 (G.F.) from the National Institutes of Health. The antibiotic resistance analysis was supported by grant no. 138082 from Empire State Development (G.F.). This research was conducted with the support of the Biorepository and Pathology CoRE at Icahn School of Medicine at Mount Sinai (RRID: SCR_027604). The collection of samples by E.D.-N. was supported by grants from Fundação de Amparo à Pesquisa do Estado de São Paulo (FAPESP – 2018/14267-2, 2018/02972-3) and Ministério da Saúde, PRONON, Brasil (SIPAR 2500.035-167/2-15-23). This work was also supported in part through the computational resources and staff expertise provided by the Department of Scientific Computing at the Icahn School of Medicine at Mount Sinai.

## Author contributions

Y.L. and G.F. designed the methods. Y.L. performed most of experiments. M.N. performed most of the computational analysis. H.C., Y.F., Y.Z. and D.W. assisted with computational analysis. X.Z., J.B., P.D, J.R.G. W.Q. Z.G and R.B. assisted with experiments. R.B. provided biopsy samples that were used in the evaluation of uHp-seq. X.Z., and M.J.B. provided additional biopsy samples for the evaluation of uHp-seq. J.B. and R.M.P. provided biopsy samples and paired cultured isolates for the validation of uHp-seq and comparison of uHp genomes with matched cultured isolates. M.C.C., E.D.-N. and A.C. provided biopsy samples to evaluate the robustness of uHp-seq. P.D. and W.S. provided biopsy samples, matched MIC data, and Deeplex HelP results for evaluating AMR prediction, virulence factor detection, and strain typing. J.B. and A.D.K. provided string samples, matched cultured isolates, MIC data, and patient treatment records and clinical outcomes to evaluate the clinical applications of uHp-seq. M.C.C. provided *Hp*GP data to assess the conservation of core methyltransferases in Hp, and perform strain typing. Collectively, these biopsy samples were used to determine the distribution of *Hp* abundance, establish the limit of detection of uHp-seq, evaluate genome recovery, characterize *in vivo* genomic heterogeneity including assess antimicrobial resistance detection, virulence factor genotyping, and strain typing. Y.L., M.N., X.Z., J.B., P.D., E.A.M., R.B, M.C.C., W.S., A.D.K., R.M.P., M.J.B. and G.F. contributed to data interpretation. Y.L. and G.F. wrote the manuscript with input and comments from all co-authors. G.F. conceived and supervised the project.

## Ethics statement

The study was approved by the relevant Institutional Review Boards and Ethics Committees, as detailed in the Methods. Written informed consent was obtained from all participants before sample collection. All procedures involving human participants were conducted in accordance with relevant ethical regulations.

## Disclaimer

This research was supported in part by the Intramural Research Program of the National Institutes of Health (NIH). The contributions of the NIH authors are considered Works of the United States Government. The findings and conclusions presented in this paper are those of the authors and do not necessarily reflect the views of the NIH or the U.S. Department of Health and Human Services.

