## Supplementary Figures for "Culture-free *Helicobacter pylori* genomics reveals within-host virulence heterogeneity and antibiotic resistance"

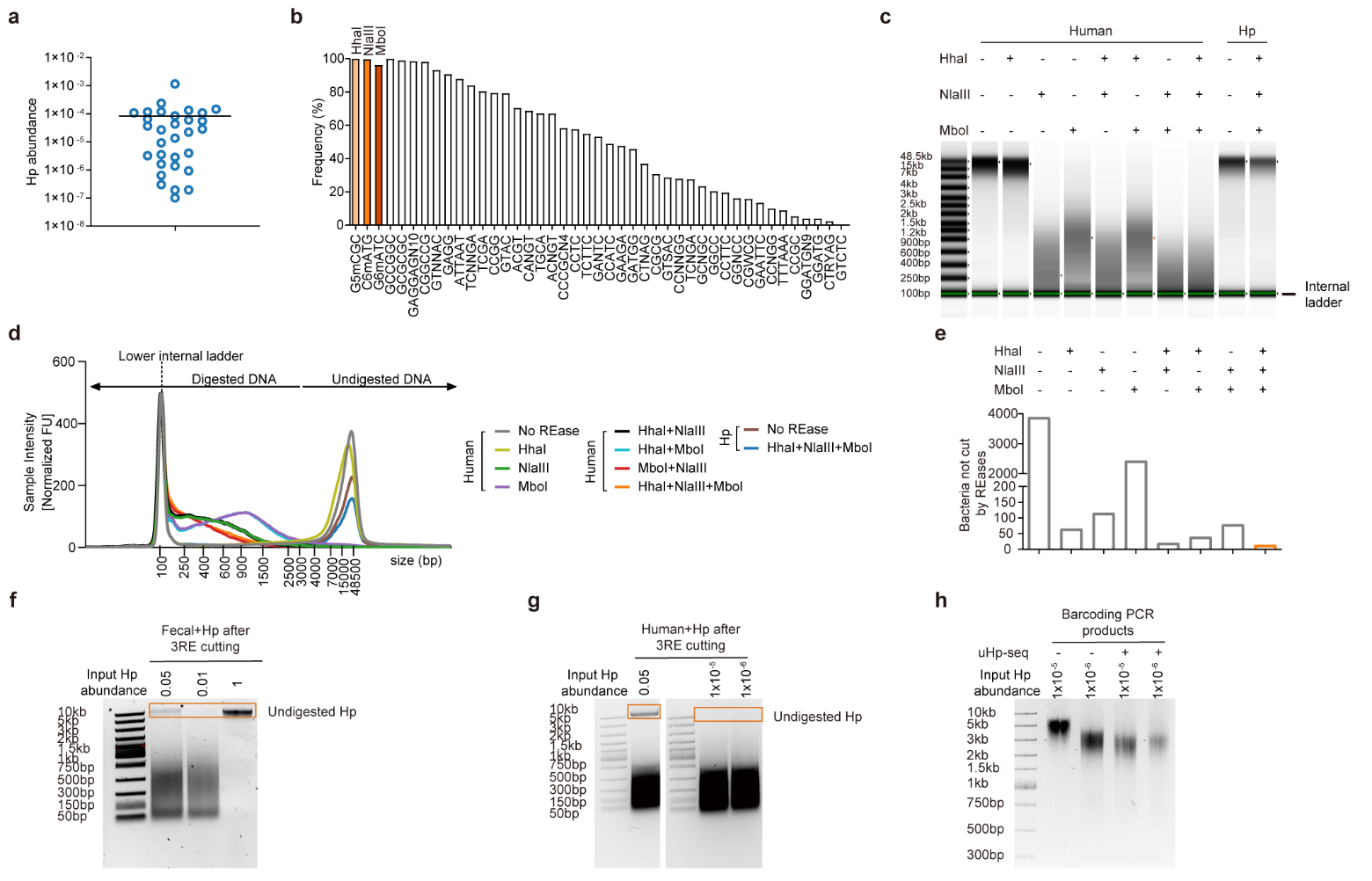

### Extended Data Figure 1. Methylation-guided REase digestion preserves long Hp DNA fragments for enrichment

**a,** Hp read abundance in gastric biopsy metagenomic sequencing datasets from publicly available studies (n=33). **b,** Frequency of all type II methyltransferases (MTases) across *Hp* genomes. Three MTases, M.HpyAVIII, M.HpyAI, and M.HpyAIII, are present in nearly all strains. Their corresponding methylation motifs are listed above, highlighting three candidate methylation-sensitive restriction endonucleases (REases) suitable for selective digestion. **c,** The electrophoresis results of DNA via TapeStation before and after indicated REases cutting separately or in combination (200ng DNA input). The bottom bands are an internal ladder. NlaIII and MboI digest human DNA well but not HhaI due to high levels of methylation at CpG islands. The combination of the chosen three REases efficiently digests human DNA but not Hp DNA. **d,** TapeStation profiles before and after REase digestion (200ng DNA input). The combination of three REases cleaves the largest proportion of non-Hp DNA into <3kb fragments while leaving methylation-protected Hp DNA as long. **e,** *In silico* digestion analysis of ~4,000 bacterial species shows that the combination of three indicated REases cleaves the largest proportion of non-Hp bacterial genomes. The analysis assessed the presence of three conserved methyltransferase recognition motifs (G^5mCGC, C^6mATG, and G^6mATC) in each bacterial genome. Genomes containing these methylation motifs were predicted to be protected from digestion by the corresponding REases, whereas genomes lacking one or more motifs were predicted to be susceptible to cleavage. **f,** Electrophoresis results of three-REase digestion applied to fecal DNA mixed with the indicated ratios of Hp DNA mocks via agarose gel (500ng DNA input). **g,** Pilot experiments using human–Hp DNA mocks demonstrate that after digestion with 3 REases, long Hp fragments are no longer detectable on conventional electrophoresis gels when the bacterial load is low (e.g., 1.0x10^-5^ and 1.0x10^-6^) (3ug DNA input). **h,** Transposase-based barcoding PCR when highly sensitive automated long-fragment size selection (e.g., BluePippin) is applied to isolate protected Hp DNA after REases cutting.

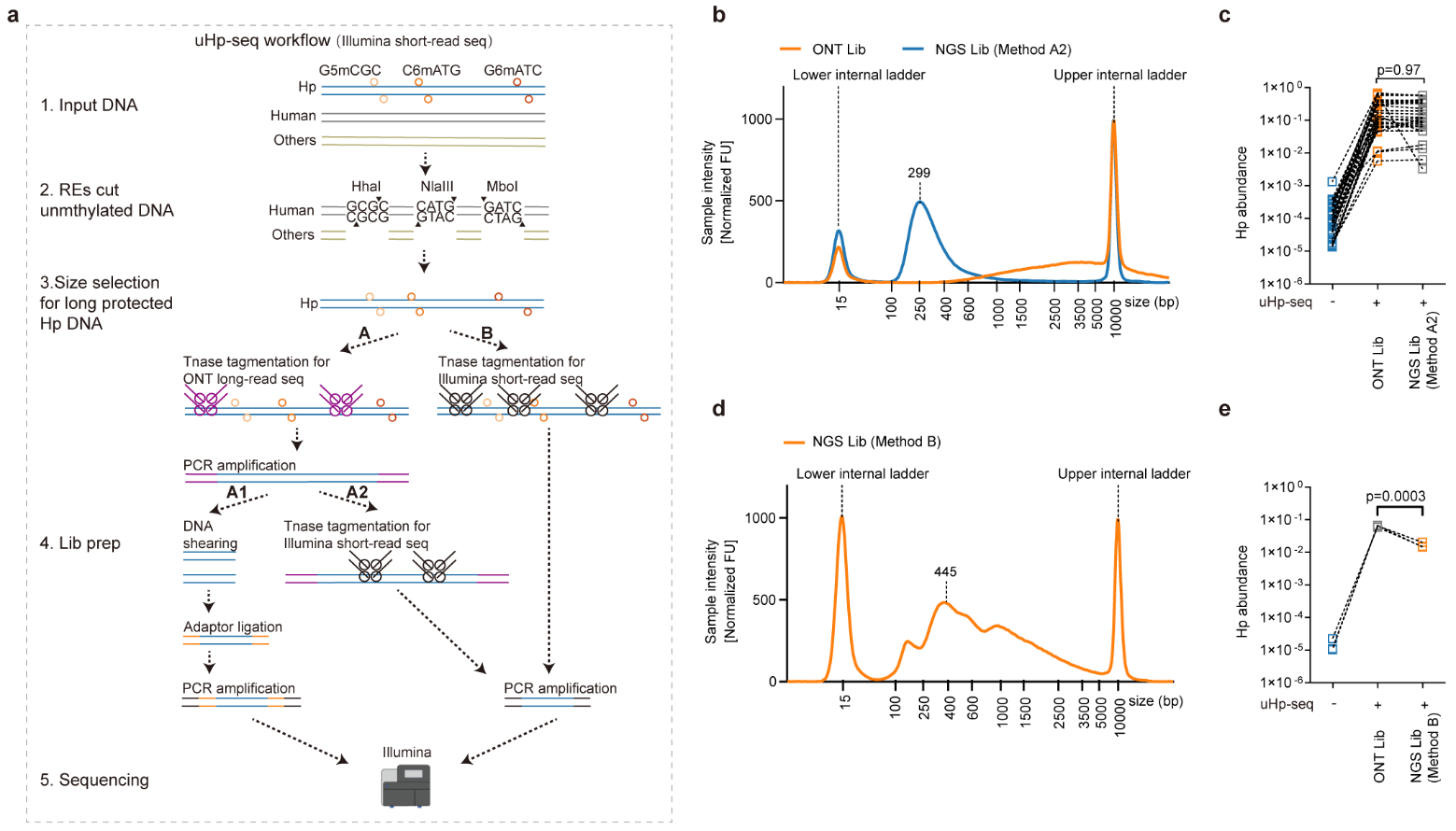

### Extended Data Figure2. Adaptation of uHp-seq for short-read sequencing

**a,** Workflow for short-read library construction in the uHp-seq protocol. Restriction enzyme digestion and size selection were performed as in the long-read uHp-seq workflow. Following size selection, short-read libraries were generated using two alternative strategies. Strategy A utilized the Tn5 tagmentation step from the long-read workflow, followed by PCR amplification. The amplified products were then either subjected to DNA fragmentation, adaptor ligation, and a second round of PCR amplification (A1), or processed through an additional round of Tn5 tagmentation optimized for short-read sequencing (A2). Strategy B bypassed the initial long-read tagmentation step and directly applied the short-read Tn5 tagmentation workflow, as described for A2. **b,** Read length distribution of uHp-seq ONT long-read libraries and NGS short-read libraries prepared using Method A2. **c,** Comparable Hp abundance observed in ONT long-read and NGS short-read libraries (constructed with method A2) following uHp-seq enrichment (n=32). **d,** Read length distribution of NGS short-read libraries prepared using Method B. **e,** Hp abundance before and after uHp-seq enrichment using Method B (n=3).

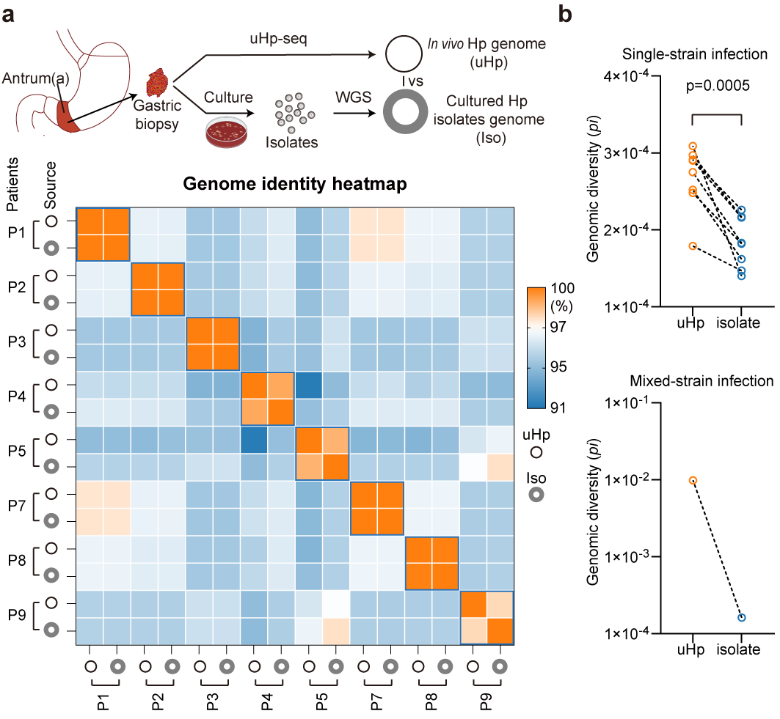

### Extended Data Figure 3. Validation of uHp-seq genome reconstruction and recovery of within-sample genetic diversity

**a,** Validation of biopsy-derived genome reconstruction. Across eight matched biopsy–isolate pairs acquired from antrum biopsies, uHp-seq genomes (uHp) showed 99.6% nucleotide identity to paired cultured isolates (Iso). **b,** Genomic diversity of nine corpus uHp samples and paired cultured isolates, including eight single-strain infections and one mixed-strain infection, measured by nucleotide diversity (*pi*) from Illumina sequencing data. Higher *pi* values indicate greater genetic diversity. Illumina data were unavailable for the paired antrum samples shown in (a).

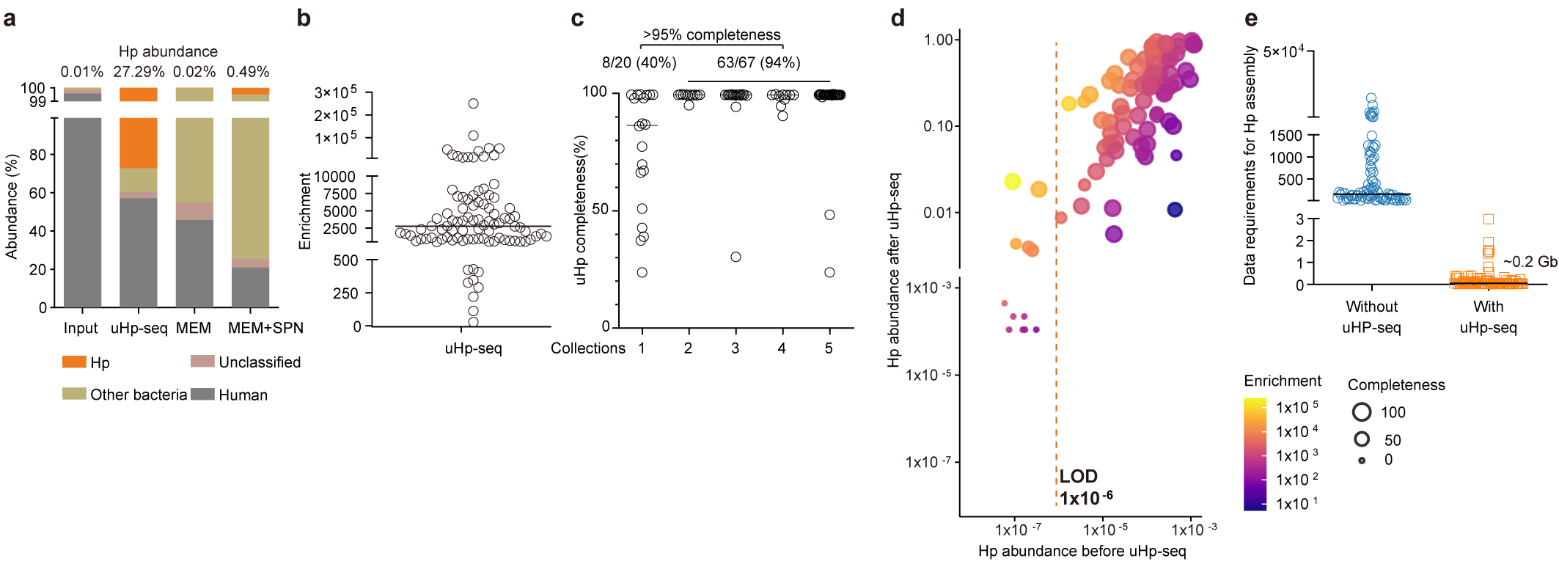

### Extended Data Figure 4. Performance evaluation of uHp-seq

**a,** Comparison of host DNA depletion methods on fresh gastric biopsies. uHp-seq achieves substantially higher Hp enrichment than MEM or MEM combined with saponin (SPN). **b,** Enrichment of Hp DNA across a large-scale clinical biopsy set processed using uHp-seq, showing robust performance across specimens (n=94). **c,** uHp genome completeness distribution across different collections (DNA samples from Collection1 had been stored for over 20 years at -80 ℃). **d,** Large-scale evaluation of uHp-seq across 94 Hp positive gastric biopsy samples. uHp-seq achieved an average ~2,500-fold enrichment and recovered high-completeness genomes from 91% of samples with input Hp abundance ≥10⁻⁶. **e,** A sequencing data output required data are ~0.2 Gb is required achieve ≥10× coverage of the Hp genome from gastric biopsies with uHp-seq. The center line in each dot plot indicates the median.

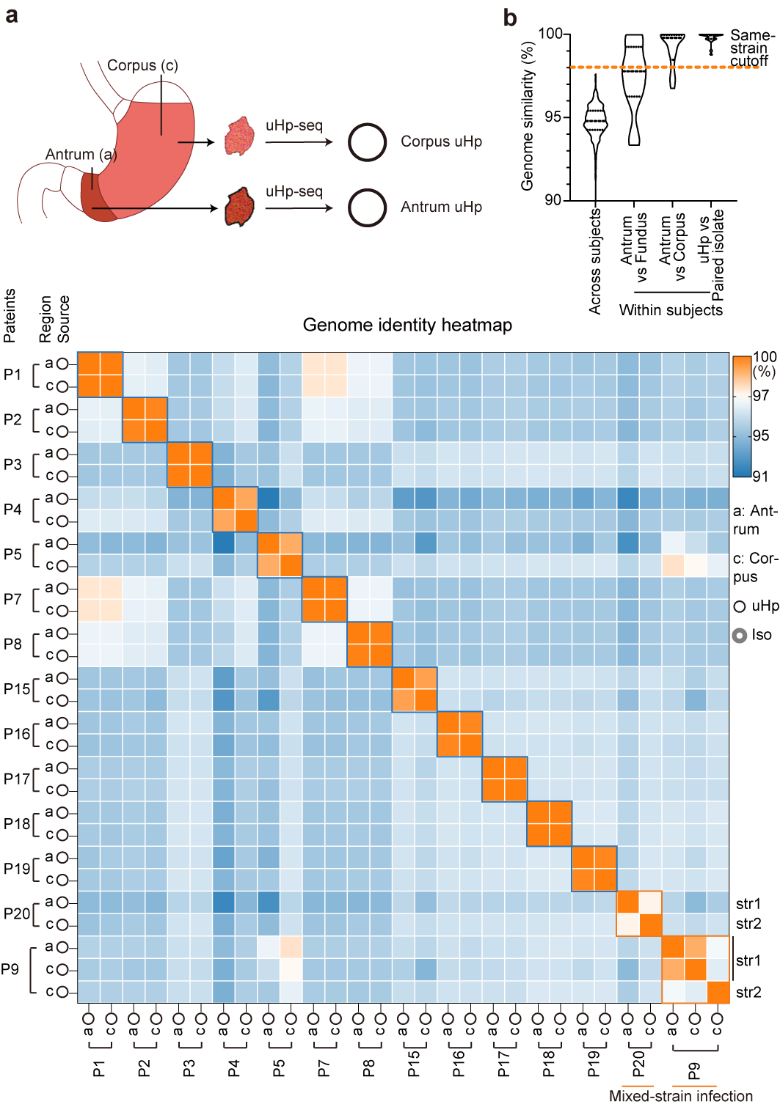

### Extended Data Figure 5.  Compared with antrum–fundus pairs, lower genomic variation between antrum uHp and paired corpus uHp

**a**, Average nucleotide identity between uHp genomes recovered from antrum and paired corpus samples. Mixed-strain infections were identified in patient P9 and P20. **b**, Distribution of ANI values among various Hp genomes. The cutoff for defining the same strain was ANI >98%.

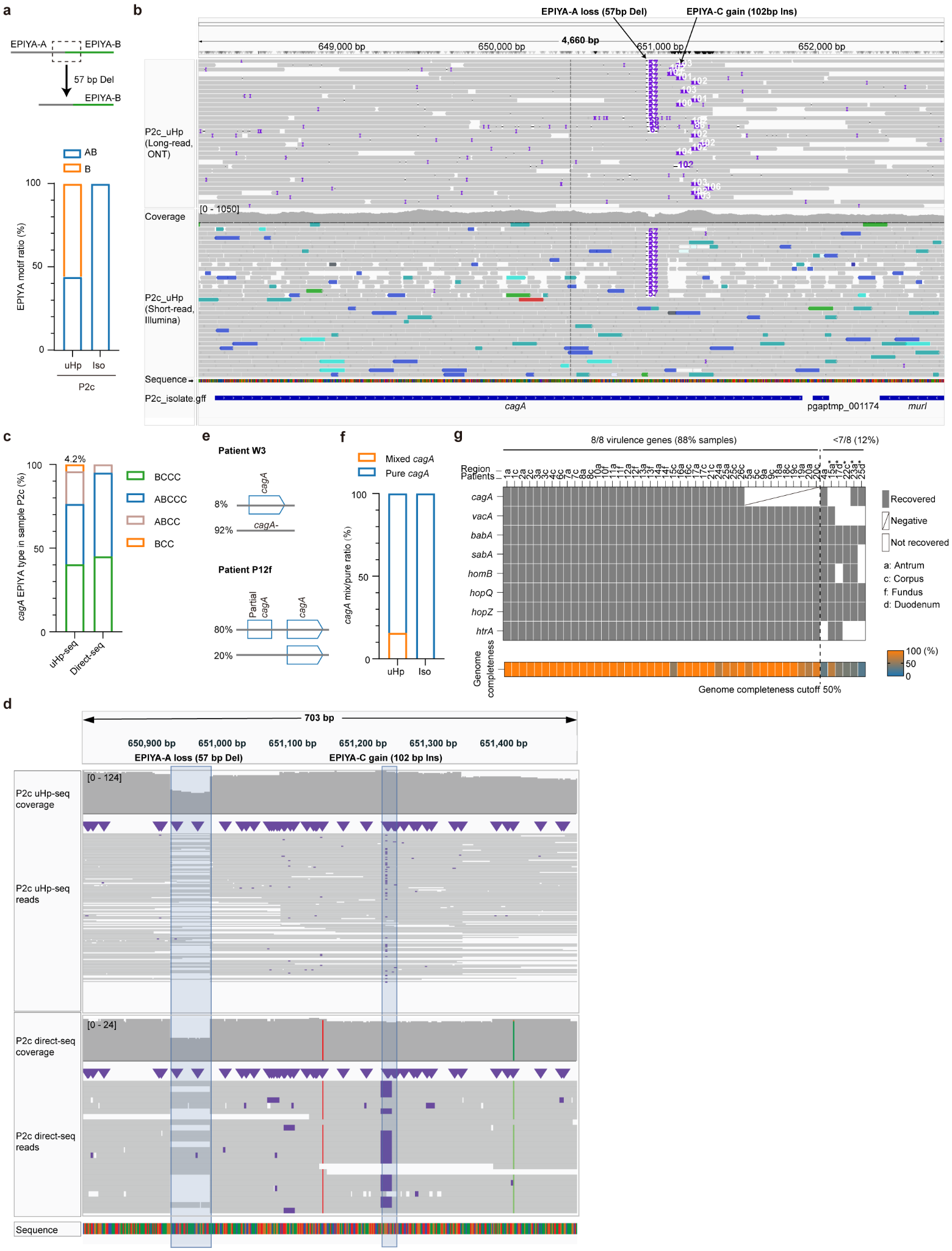

### Extended Data Figure 6. Additional virulence genes and variants uHp-seq recovered

**a,** A 57 bp deletion in *cagA* converts an EPIYA-AB motif into a novel EPIYA-B motif, resulting in the coexistence of EPIYA-AB and EPIYA-B forms in uHp genomes. This variant was not detected in the paired culture isolates. **b,** Integrative Genomics Viewer (IGV) visualization of long- and short-read mapping of P2c uHp at the *cagA* locus. The 57 bp deletion and 102 bp insertion are highlighted in purple; the 102 bp insertion cannot be resolved by short-read sequencing. **c,** *cagA* subtypes identified by uHp-seq and deep long-read sequencing (~110 Gb) of native genomic DNA from sample P2c (direct-seq), without Hp enrichment or DNA amplification. **d,** IGV visualization of reads generated by uHp-seq and direct-seq mapped to the EPIYA region of the *cagA* locus in sample P2c. **e,** Two additional patterns of *cagA* heterogeneity identified by uHp-seq. W2 harbored a mixture of *cagA*-positive (*cagA*+) and *cagA*-negative (*cagA*-) strains, whereas 80% Hp harbored two *cagA* copies in P12f, with the second copy being truncated. **f,** Proportions of mixed and pure *cagA* types identified in uHp genomes and paired culture-based isolates. **g,** Recovery of virulence-associated genes by uHp-seq across gastric biopsy samples with known anatomical sampling sites. The lower panel shows the corresponding genome completeness of each sample. A genome completeness threshold of 50% was sufficient to recover all eight virulence genes. Of the six samples in which fewer than eight genes were recovered, five were long-term stored samples from Collection 1, indicated by asterisks.

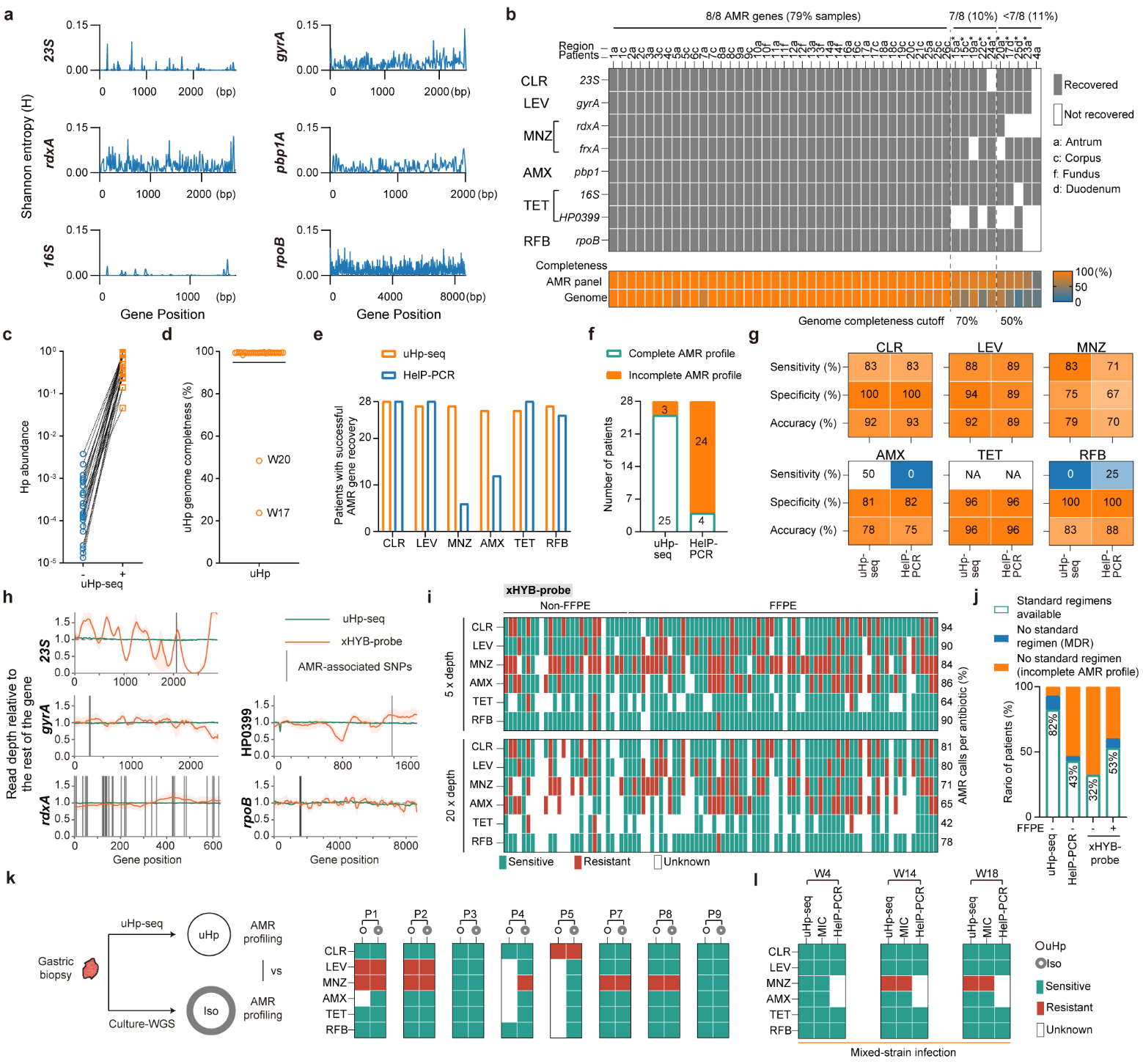

### Extended Data Figure 7. Genome-wide recovery of antibiotic resistance determinants by uHp-seq

**a,** Sequence diversity of antibiotic resistance–associated genes across Hp strains, assessed using Shannon entropy. Among the six commonly used resistance-determining genes (excluding the 16S and 23S rRNA genes), *gyrA*, *pbp1A*, *rdxA*, and *rpoB* exhibited high sequence diversity, highlighting the challenges of designing universal PCR primers capable of reliably amplifying these targets across diverse Hp strains. **b,** Recovery of eight commonly used antibiotic resistance–associated genes across all biopsy samples following uHp-seq. The lower panel shows the number of recovered resistance genes and the corresponding genome completeness for each sample. All eight resistance-associated genes were consistently recovered when genome completeness exceeded 70%, whereas at least seven of the eight genes were recovered when genome completeness exceeded 50%, demonstrating the robustness of uHp-seq for comprehensive antimicrobial resistance profiling. Of the ten samples in which fewer than eight genes were recovered, nine were long-term stored samples from Collection 1, indicated by asterisks. **c**, uHp-seq effectively enriches Hp from gastric biopsy samples in the validation cohort (n=28). **d,** uHp genome completeness estimated by CheckM (n=28). **e,** The number of patients for whom genes associated with resistance to each indicated antibiotic were successfully recovered by uHp-seq or Deeplex HelP (HelP-PCR). **f,** The number of patients with complete AMR profiles determined by uHp-seq or HelP-PCR. A complete AMR profile was defined as successful determination of resistance to all six antibiotics: CLR, LEV, MNZ, AMX, TET, and RFB. **g,** Comparison of AMR detection performance between the commercial PCR-targeted methods HelP-PCR and genome-wide resistance profiling uHp-seq. Samples with multiple-strain infection or failed AMR calls were excluded. The performance of both methods was assessed using MIC as the reference standard. NA, not applicable because no TET-resistant isolates were available for sensitivity calculation (n=28). **h,** Relative read depth across the indicated AMR-associated genes obtained by uHp-seq in the validation cohort or by Qiagen xHYB (xHYB-probe) probe-based hybridization capture in a previously published study. Vertical lines indicate the positions of AMR-associated SNPs. **i,** AMR profiles determined by XHYB-probe using minimum read-depth thresholds of 5× or 20× for SNP calling. **j,** Number of patients with an available standard treatment regimen or without an available regimen due to multidrug resistance (MDR) or incomplete AMR profiles, as determined by uHp-seq, HelP-PCR, or XHYB-probe. uHp-seq and HelP-PCR were evaluated using the same validation cohort, whereas XHYB-probe data were obtained from the published study described in **h**. A minimum read depth of 20× was required for SNP calling. **k,** Comparison of AMR profiles between paired uHp genomes and cultured isolates recovered from gastric corpus. Unknown AMR calls in P1 and P5 resulted from insufficient sequencing coverage of the corresponding genes, whereas those in P4 were attributable to lower genome completeness. **l,** Comparison of AMR profiles between uHp-seq, MIC and HelP-PCR in mixed-strain infections. Missed AMR calls (unknown) due to PCR failure in HelP-PCR.

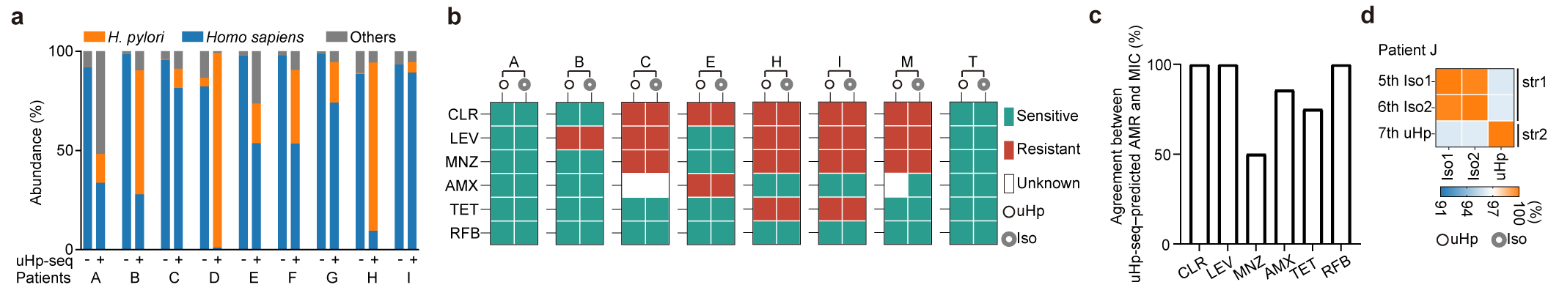

### Extended Data Figure 8. Host DNA depletion, genome recovery, and AMR profiling of string-derived Hp

**a,** Read composition of gastric string samples before and after uHp-seq. uHp-seq efficiently depleted host-derived DNA and substantially enriched Hp reads. (Only the nine indicated samples were sequenced before uHp-seq; the remaining samples were not sequenced before uHp-seq due to insufficient DNA.) **b,** Comparison of antimicrobial resistance (AMR) profiles between string-derived uHp genomes and their paired cultured isolates. **c,** Concordance between uHp-seq, predicted AMR profiles and phenotypic antimicrobial susceptibility determined by MIC testing of paired isolates. **d,** Average nucleotide identity between uHp genomes recovered from gastric string samples and Hp isolates, all obtained from patient J. Isolate 1 (Iso1) and isolate 2 (Iso2) were cultured during the fifth and sixth Hp treatment courses, respectively, while the string samples were collected during the seventh treatment course and processed by uHp-seq.

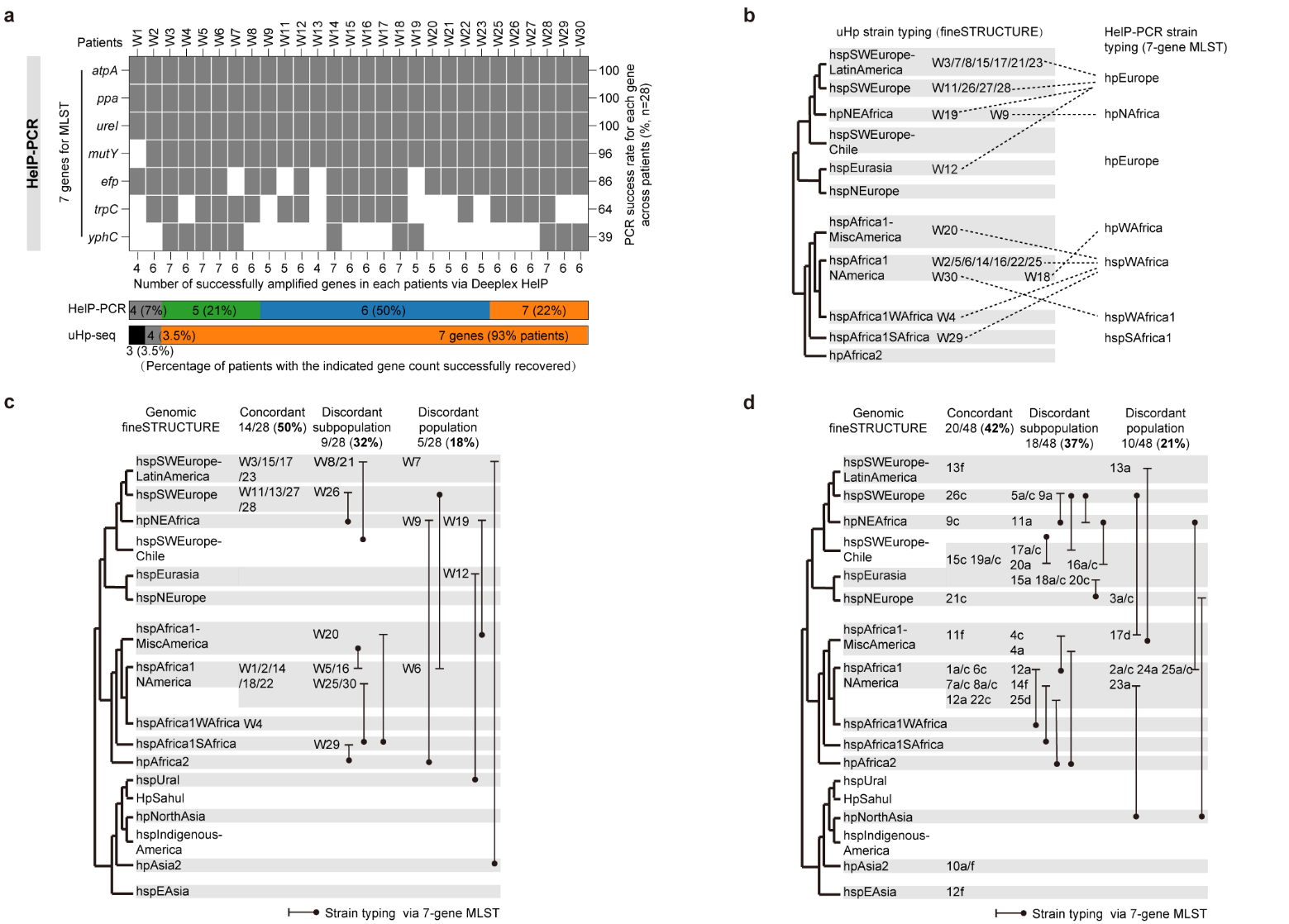

Extended Data Figure 9. High-Resolution Hp strain typing via uHp-seq

**a,** Recovery of the seven housekeeping genes required for MLST strain typing by PCR-based Deeplex HelP (HelP-PCR) and uHp-seq. Complete recovery of all seven genes was achieved in 22% of samples using HelP-PCR, compared with 93% using uHp-seq. **b,** Comparison of Hp strain assignments generated by the uHp-seq and HelP-PCR pipelines. The two methods use different Hp population classification schemes. **c–d,** Comparison of Hp strain assignments using the same population classification system of two cohorts. Genome-wide fineSTRUCTURE analysis enabled by uHp-seq was compared with conventional seven-gene MLST (performed *in silico*). Genome-wide strain typing provided more refined population assignments relevant to virulence stratification and identified discordant classifications relative to MLST. Samples with mixed-strain infection were assigned according to the dominant strain.
